# The plateau in liver disease mortality in Germany: contrasting trends by age, region and ICD-10 code, 2000–2024

**DOI:** 10.64898/2026.11.16.26368157

**Authors:** Miriam Kumpf, Era Stambollxhiu, Eva Wende, Nicolas Lenhart, Markus Graf, Sebastian Ziegelmayer, Marcus Makowski, Keno Bressem, Lisa Adams

**Affiliations:** Department of Diagnostic and Interventional Radiology, School of Medicine and Health, Technical University of Munich, Munich, Germany; National Center for Tumor Diseases West, Essen, Germany; Institute of Artificial Intelligence in Medicine, University Hospital Essen, Essen, Germany; Department of Diagnostic and Interventional Radiology and Neuroradiology, University Hospital Essen, Essen, Germany

## Abstract

**Background:** Cirrhosis is often diagnosed only at decompensation, making identification of high-risk groups important for prevention and early detection. Chronic liver disease mortality has fallen in southern Europe and France since 2010, but not in Germany. Whether the German plateau reflects birth-cohort alcohol exposure, metabolic liver disease or certification practice is unknown.

**Methods:** Deaths by ICD-10 code, sex and age group were extracted from the WHO Mortality Database for 16 European countries (2000–2022), with Eurostat rates (2011–2023), German state-level mortality (1998–2024) and hospital diagnoses (2000–2023). Registry and Global Burden of Disease Study (GBD) 2023 estimates were summarised for ages 45–64, 65–74 and 75 years and over, with registry rates standardised to the European Standard Population 2013.

**Findings:** Germany’s age-standardised chronic liver disease mortality was 16.6 per 100 000 in 2011 and 16.0 in 2023, against declines of 21–37% in France, Italy and Spain. Between 2010 and 2022, alcoholic liver disease mortality at ages 45–64 fell by 17% in men and 25% in women, whereas fibrosis and cirrhosis (K74) mortality at ages 65–74 rose by 27% and 28%, with larger K70–K77 mortality increases in eastern than western Germany. Hospitalisation rates showed similar age-specific patterns, while in-hospital case fatality declined. Fewer than 0.5% of liver deaths were certified as fatty liver. GBD 2023 showed a similar age-specific increase but attributed rising mortality to all five modelled aetiologies.

**Interpretation:** The German plateau reflects continued decline below age 65 and rising mortality at ages 65–74, compatible with cohorts born 1945–59 reaching this age band, particularly in eastern Germany. Consumption records and surveys indicate differing regional patterns of alcohol exposure. Much of the cirrhosis in these cohorts is certified without aetiology, and GBD’s aetiological estimates cannot be verified in registration data.

**Funding:** None.

**Research in context:** 

**Evidence before this study:** PubMed was searched without language restriction from inception to October 15, 2026, for registry-based analyses of cirrhosis or chronic liver disease mortality in Germany or in European comparisons, and for comparisons of GBD estimates with vital registration, using combinations of the terms cirrhosis, liver disease, alcohol-related liver disease, mortality, Germany, Europe, age-period-cohort, Global Burden of Disease and vital registration. Registry analyses have documented an east-to-west gradient of cirrhosis mortality across Europe and, in Germany, period and birth-cohort effects on alcohol-related mortality, with lower risks in later-born cohorts. A nationwide hospital study reported rising admissions for cirrhosis with falling in-hospital mortality. GBD publications acknowledge that ICD coding cannot reliably distinguish cirrhosis aetiologies, which are assigned using statistical models. We found no previous study combining detailed ICD-10 mortality analyses, age-specific European comparison, German federal-state trends and comparison with GBD aetiological estimates.

**Added value of this study:** We compared GBD 2023 estimates with European mortality registration data and examined German mortality by detailed ICD-10 code, age, sex and federal state, alongside hospital diagnosis statistics. The German plateau reflects rising mortality at 65–74 years, especially in eastern Germany, while mortality continues to decline at 45–64 years. The increase was concentrated in deaths coded as unspecified cirrhosis (K74.6) and accompanied by rising cirrhosis hospitalisation rates. At the same time, in-hospital case fatality and admissions for chronic viral hepatitis declined. GBD identified a similar age-specific increase but estimated rising mortality for all five aetiological sub-causes despite their differing representation in routine data.

**Implications of all the available evidence:** The absence of a decline in cirrhosis mortality in Germany is consistent with older birth cohorts with high cumulative alcohol exposure, although the aetiology of cirrhosis is often unspecified. Differences in alcohol consumption patterns, obesity and conditions after reunification may contribute to higher mortality in parts of eastern Germany. The findings support earlier detection and improved treatment access for older adults, age-specific surveillance in Germany and other European countries, and cautious interpretation of modelled aetiological fractions in countries without multiple-cause mortality data.

## Introduction

Cirrhosis from alcohol, viral hepatitis or metabolic disease is often diagnosed only at decompensation, so identifying high-risk groups is the basis of prevention and early detection. ^1^ Mortality from cirrhosis has fallen in most of Western Europe since the 1970s, with the largest declines in Southern Europe. ^2^ Germany is an exception in that its age-standardised death rate for chronic liver disease declined through the 2000s but has not fallen since about 2010. In the Global Burden of Disease Study 2023 (GBD 2023) the German point estimate for 2023 lies above that for 2010. ^3^

This plateau could reflect birth cohorts with high cumulative alcohol exposure reaching older ages, as age-period-cohort analyses of alcohol-related mortality in Germany suggest for cohorts born before 1960. ^4,5^ It could also reflect a rising contribution from metabolic liver disease, estimated by GBD using modelled aetiological fractions. ^6^ A French hospital-based study reported declining liver-related mortality associated with viral hepatitis and increasing mortality associated with metabolic syndrome. ^7^ Certification practice could also affect cause-specific trends, because German mortality statistics record a single underlying cause and often leave the aetiology of cirrhosis unspecified. ^8^ These explanations predict different patterns by age, region and ICD-10 code but have not been examined together using detailed German registration data.

In countries with complete vital registration, GBD estimates draw heavily on registry data but also redistribute ill-defined causes and use statistical models to estimate cause-specific mortality and cirrho-sis aetiologies. ^6,9^ Whether the German plateau and its GBD-estimated aetiologies are consistent with registration data has not been examined.

This study compares chronic liver disease mortality in Germany with 15 European countries using WHO Mortality Database and Eurostat data, examines German mortality by age, sex, region and ICD-10 code alongside hospital diagnosis statistics, and compares registry trends with GBD 2023 estimates. The aim was to characterise the German plateau and assess which explanations are supported by registration data.

## Methods

### Study design and data sources

This descriptive study used ICD-10-coded deaths by age and sex from the WHO Mortality Database (February 2026 release) for 16 European countries from 2000 onwards. ^10^ Age-band analyses used the nine countries with sufficient counts (Austria, Czechia, France, Germany, Italy, the Netherlands, Poland, Spain and the UK). Eurostat supplied published age-standardised death rates for its causes-of-death shortlist (2011–2023, 36 countries). ^11^ Eurostat mid-year populations served as denominators, except for available UK observations after 2018, for which WHO population data were used (appendix table S1). ^12^ German mortality data by federal state (Land) and for eastern and western Germany came from the Federal Statistical Office through the GBE-Bund health reporting system. ^13^ These comprise deaths by three-character cause, Land, sex and age group (under 65 and 65 years and over, 1998–2024), and deaths by ICD-10 block and 21 age groups for the 16 Länder and the eastern and western aggregates (1998–2024). Hospital diagnosis statistics were also obtained by place of residence, age group and three- or four-character main diagnosis with in-hospital deaths (2000–2023). GBD 2023 estimates for cirrhosis and other chronic liver diseases and the five modelled aetiologies (alcohol, hepatitis B, hepatitis C, non-alcoholic fatty liver disease and other causes) were retrieved through the IHME data connector as age-standardised rates with 95% uncertainty intervals for 29 countries and regions (2010–2023) and as five-year age-specific rates for the nine selected countries (2000–2023). ^14^ Query parameters and file versions are listed in appendix table S1.

### Case definitions

The ICD-10 codes analysed were alcoholic liver disease (K70), fibrosis and cirrhosis of the liver (K74), including K74.6 (other and unspecified cirrhosis of liver), and other diseases of the liver (K76), including K76.0 (fatty liver). Combined categories were K70, K73 (chronic hepatitis) and K74 (Eurostat’s definition of chronic liver disease), K70–K76 (the closest registry equivalent of the GBD cause) and K70–K77 (the liver disease block in the GBE-Bund age tables). Liver cancer (C22), chronic viral hepatitis (B18) and alcohol use disorders (F10) were also analysed. Hospital analyses used alcoholic cirrhosis (K70.3), K74.6 and K76.0 as main diagnoses.

### Age harmonisation and standardisation

WHO age columns were mapped to 21 groups according to the age format of each row (appendix section S3). Deaths of unknown age were distributed proportionally across known age groups. Age-specific cells suppressed for confidentiality (Germany from 2021, fewer than three deaths) were imputed by proportionally scaling the reported age-specific counts to the complete all-ages totals (appendix section S3). Rates were directly standardised to the European Standard Population 2013 (ESP2013) and, for overall comparison with GBD, to the WHO world standard. Rates for ages 45–64, 65–74 and 75 years and over were ESP2013-weighted means of five-year rates. The same within-band weights were applied to GBD and GBE-Bund rates.

### Regional and hospital analyses

Eastern and western Germany were defined using the GBE-Bund aggregates of the new Länder excluding former East Berlin and the former federal territory including East Berlin, respectively. Comparisons between Länder used arithmetic means of the annual band rates over three years (2009–11, 2018–20 and 2022–24), and Länder with fewer than 30 deaths per year in a band were not interpreted. In-hospital case fatality was calculated as in-hospital deaths per 100 stays.

### Exposure data

Historical per-capita alcohol consumption in the GDR and FRG was obtained from Sieber and colleagues, ^15^ and obesity prevalence by Land from the Mikrozensus. ^16^ Published BGS98^17^ and GEDA ^13,18^ estimates were used to examine drinking patterns in eastern and western Germany. While GEDA 2009–12 used AUDIT-C, the data for 2014/15 are based on a different, intake-based definition ^19^, limiting direct comparability. Supplementary mortality rates for mental and behavioural disorders due to alcohol (F10) and alcoholic liver disease (K70) for 1998–2024 were obtained from GBE-Bund. Data sources, regional definitions and aggregation methods are detailed in appendix section S6.

### Validation and statistical analysis

Independently calculated ESP2013-standardised rates for K70, K73 and K74 combined were compared with corresponding Eurostat rates for the same country, year and sex (median absolute difference 0.2% for Germany and below 1% for 22 of 31 countries). GBE-Bund death counts were compared with WHO counts, which were identical up to 2020. K70 and K74 counts differed by less than 0.3% in 2021 and 2022 after rescaling (appendix section S3, table S3). Rates are reported per 100 000 with the standard population stated. Changes were calculated between the first comparison year (2011 for Eurostat and 2010 for the other sources) and the final available year (2022 for WHO data, 2023 for Eurostat, GBD and hospital data and 2024 for German deaths) unless otherwise specified, or between three-year means. Hospitalisation trends were assessed from 2010 to 2019, the last pre-pandemic year, because hospital stays subsequently declined and remained below their 2019 levels in 2023. All mortality analyses are descriptive, and no inferential tests were performed. GBD estimates are reported with their published 95% uncertainty intervals. All analysis steps were implemented using Python scripts (Python 3.13, pandas 2.3.3, matplotlib 3.10). The analysis pipeline and query parameters are described in the appendix, and scripts and the data log are available at github.com/m-kumpf/liver-mortality-germany. Reporting followed applicable STROBE recommendations. Ethical approval was not required for aggregated public data.

## Results

### Germany against Europe

In the Eurostat series for chronic liver disease (K70, K73 and K74, both sexes), Germany’s age-standardised mortality rate was 16.6 per 100 000 in 2011, 15.2 in 2019 and 16.0 in 2023 (15 004 deaths).

Between 2011 and 2023, rates declined in France (12.0 to 9.4, *−*21%), Italy (10.4 to 6.5, *−*37%), Spain (11.0 to 7.5, *−*31%), and Austria (17.1 to 14.8, *−*13%). The rate remained near 5 in the Netherlands but increased in Poland (17.7 to 20.7) and Czechia (18.5 to 19.5; figure 1A–C).

**Figure 1.**
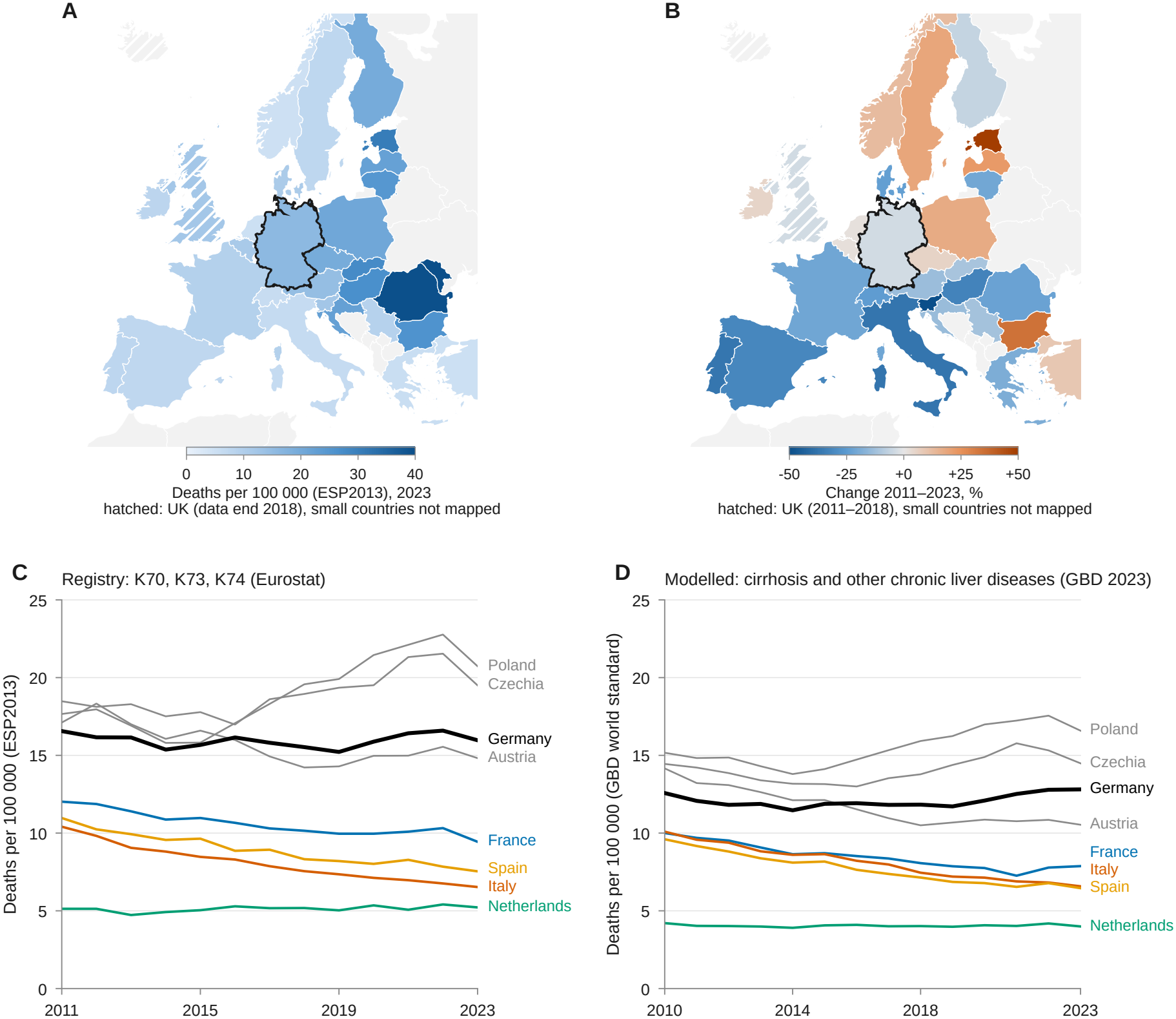
Chronic liver disease mortality in Germany and Europe. (A) Age-standardised death rate from chronic liver disease (ICD-10 K70, K73, K74, European Standard Population 2013) in 2023, both sexes, 36 countries (Eurostat). (B) Percent change in the same rate between 2011 and 2023. The UK (hatched) is shown for 2018, its last year in the Eurostat series. Iceland, Liechtenstein, Luxembourg, Malta and Cyprus are not mapped because of small numbers. (C) Annual rates 2011–2023 for Germany and the comparator countries, with Austria, Poland and Czechia in grey. (D) Modelled age-standardised death rate from cirrhosis and other chronic liver diseases, GBD 2023 (GBD world standard population), 2010–2023, for the same countries and the Western Europe region (dashed). Rates in C and D differ in case definition, standard population and redistribution of ill-defined causes. 95% uncertainty intervals for the GBD change are given in appendix table S5.

GBD 2023 showed a similar pattern. Germany’s modelled age-standardised rate was 12.6 per 100 000 in 2010 and 12.8 in 2023 (+1.9%, 95% UI *−*6.5 to +13.2), while the Western European rate fell from 10.1 to 8.7. GBD estimated declines in France (*−*21%), Italy (*−*35%) and Spain (*−*33%), and increases in the UK (+17%) and Poland (+9%; figure 1D). GBD rates exceeded registered K70–K76 mortality by approximately 15–25% in several Western European countries (mean GBD-to-registry ratio for 2015–2022 of 1.22 for Germany, 1.42 for Italy), while Czechia (1.01) and the UK (0.95) were close to parity. In Germany, the ratio ranged from 1.14 to 1.22 across the age bands (appendix figure S2).

In the German four-character data, the rate for K70–K76 (both sexes) fell from 23.8 per 100 000 in 2000 to 18.2 in 2010 and 16.8 in 2019, before rising to 18.3 in 2022 (17 198 deaths). Alcoholic liver disease mortality fell from 10.3 in 2010 to 8.4 in 2019, then rose to 9.3 in 2022, while mortality from fibrosis and cirrhosis increased from 6.5 to 7.2 between 2010 and 2022 (5464 to 6940 deaths). K74.6 accounted for approximately 98% of K74 deaths throughout the study period. Mortality registered as chronic viral hepatitis (B18) declined from 1.00 per 100 000 (854 deaths) in 2010 to 0.49 (459 deaths) in 2022, a reduction of 51%.

### Age-specific pattern in Germany

Between 2010 and 2022, mortality from alcoholic liver disease (K70) at ages 45–64 fell from 31.3 to 26.0 per 100 000 in men (*−*17%) and from 12.6 to 9.5 in women (*−*25%). At ages 65–74, however, it rose from 36.3 to 38.9 in men (+7%) and from 12.8 to 13.7 in women (+7%). At 75 years and over, rates declined in men and increased slightly in women (table 1, figure 2).

**Table 1.** Mortality from alcoholic liver disease and from fibrosis and cirrhosis in Germany by sex and age band, 2010, 2019 and 2022.

|  |  |  | 2010 |  | 2019 |  | 2022 |  | Change |
|---|---|---|---|---|---|---|---|---|---|
| Cause | Sex | Age (years) | Rate | Deaths | Rate | Deaths | Rate | Deaths | 2010–2022 |
| Alcoholic liver disease (K70) | Men | 45–64 | 31.3 | 3 480 | 24.0 | 2 994 | 26.0 | 3 224 | −17% |
|  |  | 65–74 | 36.3 | 1 605 | 35.3 | 1 431 | 38.9 | 1 702 | +7% |
|  |  | ≥75 | 21.8 | 619 | 18.5 | 742 | 19.7 | 732 | −10% |
|  | Women | 45–64 | 12.6 | 1 414 | 8.9 | 1 116 | 9.5 | 1 192 | −25% |
|  |  | 65–74 | 12.8 | 629 | 12.1 | 547 | 13.7 | 669 | +7% |
|  |  | ≥75 | 6.6 | 302 | 6.7 | 367 | 6.9 | 351 | +5% |
| Fibrosis and cirrhosis (K74) | Men | 45–64 | 10.3 | 1 112 | 10.0 | 1 252 | 10.6 | 1 349 | +3% |
|  |  | 65–74 | 23.9 | 1 067 | 27.0 | 1 074 | 30.4 | 1 325 | +27% |
|  |  | ≥75 | 36.2 | 956 | 38.8 | 1 487 | 39.4 | 1 496 | +9% |
|  | Women | 45–64 | 5.1 | 555 | 4.4 | 549 | 4.9 | 625 | −4% |
|  |  | 65–74 | 12.3 | 622 | 13.8 | 609 | 15.7 | 767 | +28% |
|  |  | ≥75 | 21.0 | 980 | 21.8 | 1 257 | 21.0 | 1 193 | 0% |
| K70 and K74 combined | Men | 45–64 | 41.6 | 4 592 | 34.0 | 4 246 | 36.6 | 4 573 | −12% |
|  |  | 65–74 | 60.2 | 2 672 | 62.3 | 2 505 | 69.3 | 3 027 | +15% |
|  |  | ≥75 | 58.0 | 1 575 | 57.3 | 2 229 | 59.1 | 2 228 | +2% |
|  | Women | 45–64 | 17.7 | 1 969 | 13.3 | 1 665 | 14.4 | 1 817 | −19% |
|  |  | 65–74 | 25.1 | 1 251 | 25.9 | 1 156 | 29.4 | 1 436 | +17% |
|  |  | ≥75 | 27.5 | 1 282 | 28.5 | 1 624 | 27.9 | 1 544 | +1% |

Rates are deaths per 100 000 standardised to the European Standard Population 2013 within the age band. Deaths are underlying-cause counts (WHO Mortality Database, Eurostat mid-year population). K74.6 (unspecified cirrhosis) accounts for 98% of K74 in every year.

**Figure 2.**
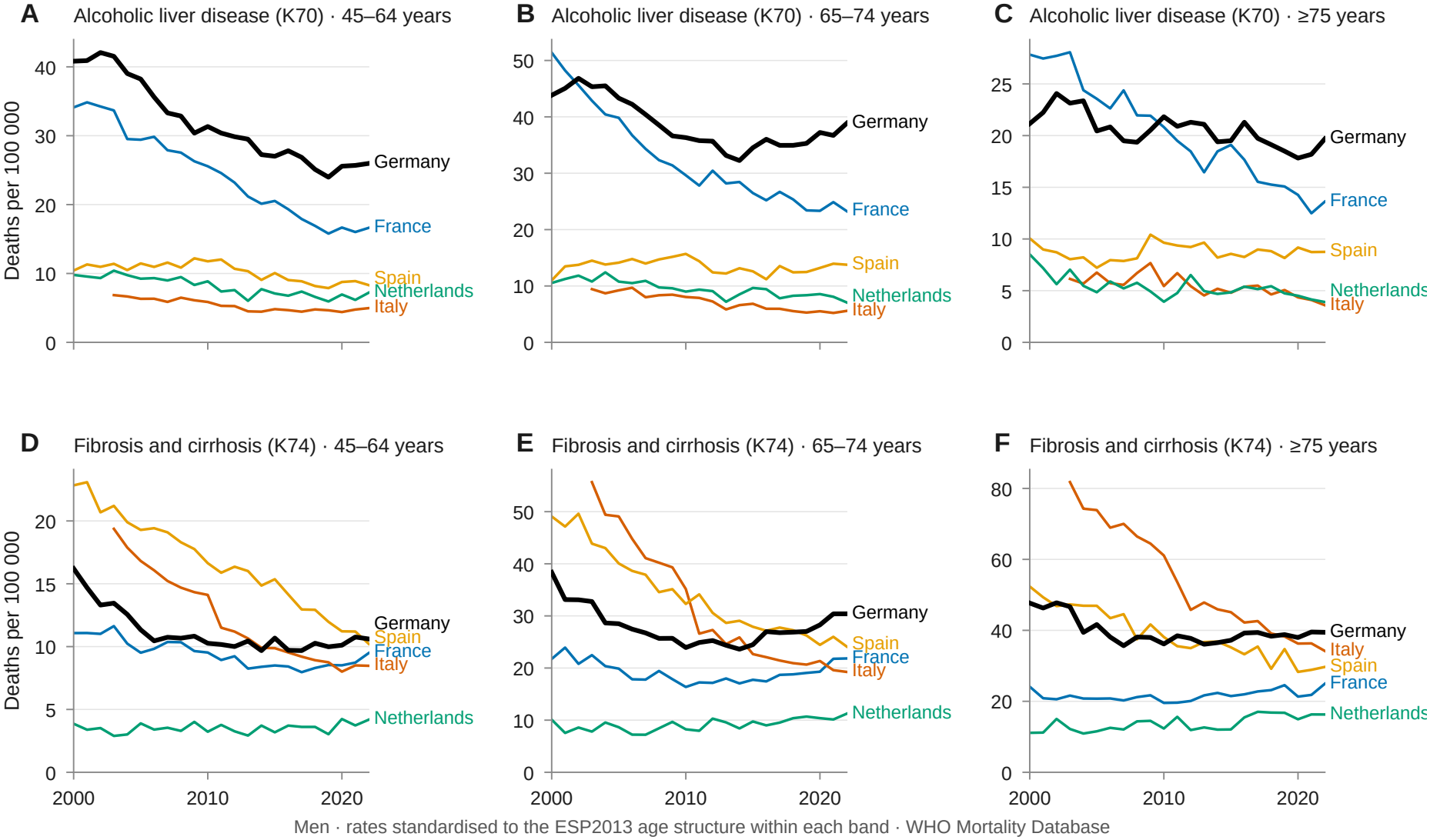
Age-specific mortality from alcoholic liver disease and fibrosis and cirrhosis in men, 2000–2022. Mortality rates per 100 000 for K70 (A–C) and K74 (D–F), at ages 45–64, 65–74 and 75 years and over, standardised to ESP2013 within each age band. Data are shown for Germany, France, Italy, Spain and the Netherlands (WHO Mortality Database, Eurostat population denominators). Corresponding results for women are shown in appendix figure S1.

Mortality from fibrosis and cirrhosis (K74) increased at ages 65–74, from 23.9 to 30.4 per 100 000 in men (+27%) and from 12.3 to 15.7 in women (+28%). Rates remained broadly stable at ages 45–64. At 75 years and over, they rose by 9% in men and remained unchanged in women. For K70 and K74 combined, mortality declined by 12% in men and 19% in women at ages 45–64, but increased by 15% and 17%, respectively, at ages 65–74.

At ages 65–74, K74 mortality also increased in France (+33% in men, +27% in women) and the Netherlands (+36%, +59%), whereas K70 and K74 mortality declined across age groups in Italy and Spain. Austria, Poland and Czechia showed declining K74 mortality alongside rising K70 mortality.

### GBD 2023 against the registry age pattern

GBD 2023 identified increasing mortality at ages 65–74 years in Germany, although the magnitude of change differed from the registry data. Between 2010 and 2022, mortality from K70–K76 in men changed by *−*12%, +18% and +6% at ages 45–64, 65–74 and 75 years and over, compared with *−*2%, +10% and *−*2% in GBD. The corresponding changes in women were *−*17%, +21% and +8% in the registry, compared with *−*8%, +12% and *−*4% in GBD. In contrast, GBD closely matched registered declines at ages 65–74 in Italy (men *−*33% versus *−*30%, women *−*50% versus *−*49%) and Spain (men *−*17% in both sources; appendix figure S2).

At ages 65–74 years, GBD estimated increases in mortality from all five aetiologies in Germany between 2010 and 2023, by 7% for alcohol, 18% for hepatitis B, 10% for hepatitis C, 17% for non-alcoholic fatty liver disease and 6% for other causes. The corresponding proportions of GBD-modelled mortality in 2023 were 48%, 7%, 15%, 18% and 12%. In registered mortality data, K74 accounted for 45% of K70 and K74 deaths combined in Germany in 2019 (38% in 2010), compared with 82% in Italy, 71% in Spain, 45% in France and 25–27% in Czechia, the UK and Poland in 2019. Deaths coded as fatty liver (K76.0) accounted for 0.1–0.4% of registered K70–K76 deaths in Germany (29 deaths in 2010 and 74 in 2022), compared with 5% in the UK in 2019.

### Eastern and western Germany

Between 2010 and 2024, mortality from liver disease (K70–K77) at ages 65–74 rose by 18% in men in eastern Germany (82.1 to 96.8 per 100 000), compared with 6% in western Germany (58.6 to 62.0). Among women, it rose from 29.7 to 37.9 in eastern Germany (+28%) and from 26.8 to 30.1 in western Germany (+12%; figure 3A, B). At ages 45–64, mortality declined by 19–20% among men in both regions and by 8% in eastern and 24% in western Germany for women. The ratio of eastern to western German mortality at ages 65–74 increased from 1.40 to 1.56 in men and from 1.11 to 1.26 in women.

**Figure 3.**
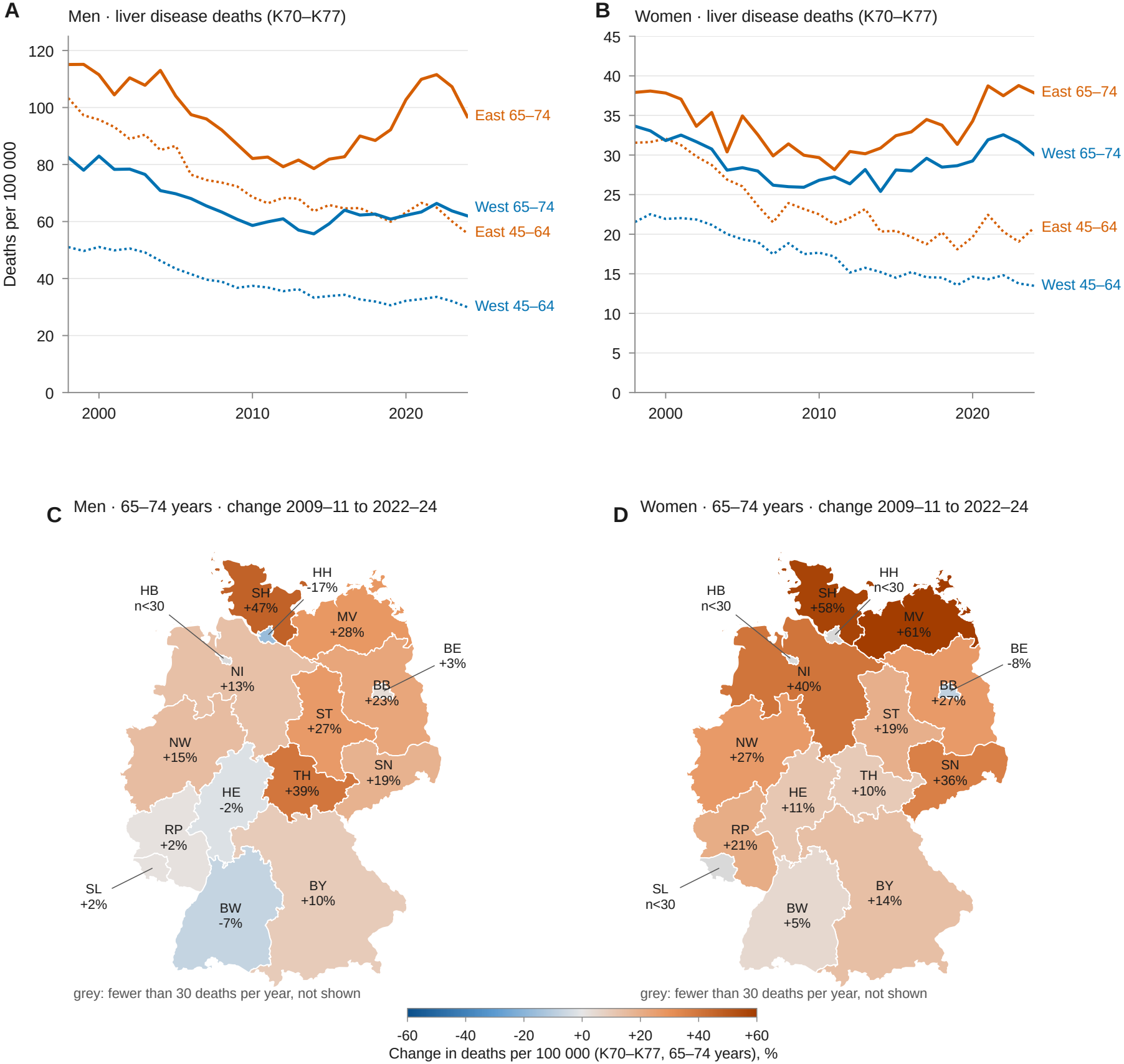
Liver disease mortality at 65–74 years within Germany. (A, B) Deaths from diseases of the liver (K70–K77) per 100 000 at 65–74 years (solid) and 45–64 years (dotted) in eastern Germany (new Länder without East Berlin) and western Germany (former federal territory with East Berlin), 1998–2024, men and women (Todesursachenstatistik, GBE-Bund). (C, D) Percent change in the 65–74-year rate between the three-year means 2009–11 and 2022–24 by Land, men and women. Länder with fewer than 30 deaths per year in this age band in 2022–24 are shown in grey because their changes are unstable. These are Bremen (23 deaths per year in men, 12 in women), Hamburg (22 in women) and Saarland (28 in women). All other Länder have at least 32 (men, Hamburg) and 50 (women, Thuringia) deaths per year.

Among men aged 65–74, mortality increased in all five eastern Länder between 2009–11 and 2022–24 (three-year means), ranging from +19% in Saxony to +39% in Thuringia (69 to 96 per 100 000). Increases also occurred in Schleswig-Holstein (+47%), North Rhine-Westphalia (+15%) and Lower Saxony (+13%), while rates declined in Hamburg (*−*17%) and Baden-Württemberg (*−*7%) and changed less elsewhere (figure 3C). Among women, the largest interpretable increases were in Mecklenburg-Western Pomerania (+61%), Schleswig-Holstein (+58%), Lower Saxony (+40%) and Saxony (+36%; figure 3D). Across the Länder, K74 accounted for 24% (Saxony-Anhalt) to 58% (Saarland) of the K70 and K74 deaths combined (all ages and sexes, 2018–20; appendix table S7).

Recorded per-capita consumption of pure alcohol was higher in the Federal Republic than in the GDR from 1960 to 1989 (11.8 vs 10.9 L in 1989), but a higher proportion of consumed ethanol came from spirits in the GDR (47% vs 18% in 1988; appendix figure S3) ^15^. The unweighted mean obesity prevalence among adults was higher in the eastern Länder in 1999 (13.1% vs 9.8%) and 2021 (16.8% vs 15.7%), although a methodological change in the Mikrozensus limits comparisons over time (appendix figure S3C). In supplementary analyses of registered alcohol-specific mortality, the eastern K70 excess was most pronounced among men aged 65 years and over. At ages 65 and over, F10 mortality increased in both regions after 2010 and was higher among women in western Germany (appendix figure S5).

### Hospital stays

Between 2010 and 2019, hospitalisation rates for unspecified cirrhosis (K74.6) at ages 65–74 increased by 28% in both eastern and western Germany (60.8 to 77.8 and 53.0 to 68.1 per 100 000, respectively), with smaller changes at ages 45–64. Rates for alcoholic cirrhosis (K70.3) increased at ages 65–74 by 14% in eastern and 7% in western Germany but declined by 10% in both regions at ages 45–64 (figure 4A). Hospitalisation rates for fatty liver (K76.0) remained low without a sustained increase. At ages 65–74, rates for alcohol use disorder (F10) rose by 29% in eastern and 4% in western Germany, while liver cancer (C22) increased by 13–20%. In contrast, hospitalisation rates for chronic viral hepatitis (B18) declined by 54–84% across all age bands and regions (figure 4B).

**Figure 4.**
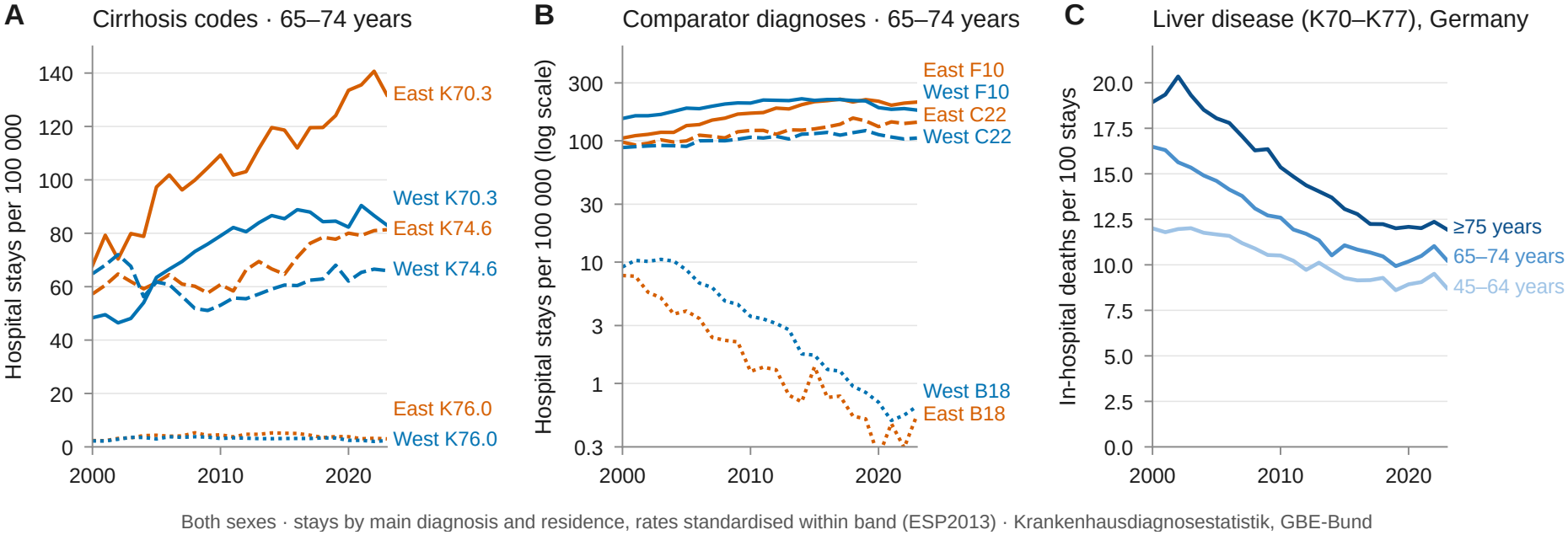
Hospital stays for liver disease, 2000–2023, both sexes. (A) Stays per 100 000 at 65–74 years with a main diagnosis of alcoholic cirrhosis (K70.3), unspecified cirrhosis (K74.6) and fatty liver (K76.0), eastern and western Germany. (B) Stays at 65–74 years for alcohol use disorders (F10), liver cancer (C22) and chronic viral hepatitis (B18). The y-axis is logarithmic. (C) In-hospital deaths per 100 stays for diseases of the liver (K70–K77) by age band, Germany. Hospital stays are counted per admission. Rates are standardised within band to the ESP2013 (Krankenhausdiagnosestatistik, GBE-Bund). Hospitalisation changes reported in the text refer to 2010–2019 (before the pandemic), the figure shows the complete series through 2023.

In-hospital case fatality for liver disease (K70–K77) at ages 65–74 declined from 16.5% in 2000 to 9.9% in 2019, followed by a slight increase to 10.3% in 2023. At ages 75 years and over, it declined from 18.9% to 12.0% between 2000 and 2023 (figure 4C).

## Discussion

The registry data show that Germany’s liver disease mortality plateau reflects opposing age-specific trends. Mortality declined at ages 45–64 but increased at 65–74, particularly for unspecified cirrhosis (K74.6). The increase was most pronounced in eastern Germany but also occurred in parts of northern and western Germany. Hospitalisation rates showed a similar age-specific pattern. GBD 2023 identified an increase in the same age group but estimated rising mortality for all five modelled aetiologies.

The observed age and regional patterns are consistent with the ageing of birth cohorts with high cumulative alcohol exposure. Age-period-cohort analyses of alcohol-attributable mortality in Germany found lower risks in cohorts born after 1945–49 in western Germany and after 1955–59 in eastern Germany, ^5^ and a six-country analysis found lower risks in cohorts born in the 1960s and later. ^4^ A previous analysis identified persistent excess mortality among men from East Germany born around 1950–70 and a renewed increase in chronic liver disease mortality at ages 60–74 after 2013. ^20^ Similar increases in alcohol-specific mortality among older adults have been reported across Europe. ^21^

Per-capita alcohol consumption was higher in the FRG than in the GDR before reunification, but spirits accounted for a substantially greater share in the GDR. In BGS98, adjusted mean alcohol intake was higher among eastern than western German men (19.2 vs 16.8 g/day), ^17^ and GEDA 2009–12 reported more risky drinking among eastern men aged 45–64 (AUDIT-C, 39.2% vs 29.1%; appendix table S9 and figure S4) ^13,18^. These differences in drinking behaviour may have contributed to the regional mortality pattern. Higher obesity prevalence in eastern Germany also suggests a possible contribution from metabolic risk factors. ^16^ Socioeconomic conditions after reunification may also have contributed. ^20^

The mortality increase at ages 65–74 was accompanied by rising hospitalisation rates for alcoholic and unspecified cirrhosis, despite declining in-hospital case fatality. A previous nationwide German study likewise found increasing cirrhosis admissions and declining in-hospital mortality (from 11.6% to 9.5%) between 2005 and 2018, despite an increase in median age from 63 to 65 years. ^22^

The agreement between hospitalisation and mortality trends argues against an explanation based solely on changes in death certification. K74.6 had a positive predictive value of 91% for chart-confirmed cirrhosis in a Swedish hospital and specialist outpatient registry, ^23^ but has not been validated as a death code in Germany. German cause-of-death statistics record a single underlying cause, and the aetiology of cirrhosis is frequently unspecified. ^8^ Differences in the proportion of deaths recorded under K70 versus K74 across countries and federal states may reflect both underlying disease patterns and certification practice. ^24^ The rise in K74.6 mortality at ages 65–74 therefore indicates an increasing burden of cirrhosis whose aetiology cannot be determined from death-registration data.

Historical differences in the burden of viral hepatitis may have contributed to the divergent cirrhosis mortality trends in Germany and southern Europe. In 2019, an estimated 459 000 people were living with chronic HCV infection in Italy, compared with 197 000 in Germany. ^25^ In a Veneto study, the three-year mortality risk among patients with HCV-related cirrhosis nearly halved between the 2013 and 2020 cohorts, while it remained stable among patients with alcohol-related cirrhosis. ^26^ French data likewise reported declining viral-(*−*42%) and alcohol-related (*−*17%) liver mortality between 2011 and 2023, but increasing mortality associated with metabolic syndrome. ^7^ In our registry analysis, however, K74 mortality at ages 65–74 increased in both France and the Netherlands, showing that similar age-specific trends can occur despite differing overall mortality trajectories. A recent Polish registry analysis likewise identified increasing K70/K74 mortality at ages 65–74 after 2015. ^27^

GBD mortality estimates exceeded registered K70–K76 mortality by 15–25% in several Western European countries. GBD redistributes ill-defined causes to specific causes, which increased estimated cirrhosis deaths by 26% in GBD 2010. ^28^ GBD estimates overall cirrhosis mortality from cause-of-death data and statistical models, then allocates it among five aetiologies using modelled proportions informed by alcohol consumption, hepatitis B and C prevalence, and obesity. ^3,6^ Because the five aetiologies share covariates and modelling assumptions, their estimated trends are not independent of each other. German death-registration data cannot independently validate the aetiological proportions estimated by GBD.

Metabolic liver disease may contribute to rising cirrhosis at older ages, but German death-registration data cannot quantify its contribution. Fewer than 0.5% of deaths were coded as fatty liver (K76.0), although a nationwide hospital study found a fourfold increase in NAFLD-associated cirrhosis admissions between 2005 and 2018. ^22^ German claims data identified advanced liver disease in over a quarter of patients coded for metabolic steatohepatitis (K75.8). ^29^ Alcohol and metabolic risk factors often coexist, and their contributions to K74.6-coded deaths cannot be separated in these data. Mortality from chronic viral hepatitis (B18) declined by 51% between 2010 and 2022, alongside falling B18 hospitalisation rates. GBD nevertheless estimated a 10% increase in hepatitis C-related cirrhosis mortality at ages 65–74 between 2010 and 2023. These are distinct outcomes and are not directly comparable.

The temporary increase in mortality during 2020–21 is consistent with the pandemic-related changes reported in Germany, when alcohol-specific deaths rose despite fewer hospital admissions. ^30^ The subsequent partial reversal does not alter the longer-term increase at ages 65–74.

This study has limitations. German cause-of-death statistics record only one underlying cause, and the aetiology of cirrhosis is often unspecified. ^8^ Age-specific WHO mortality counts with fewer than three deaths were suppressed from 2021 and required imputation, although sensitivity analyses showed little effect on the principal K70 and K74 estimates. Hospital stays represent admissions rather than individual patients and cannot be interpreted as disease incidence. Differences in cause definitions, coding practices and age-group resolution limit comparability between data sources. No age-period-cohort model was fitted, and lifetime exposure to alcohol in the relevant cohorts was not measured. Nevertheless, mortality and hospitalisation data show concordant age-specific trends, with increasing liver disease burden at ages 65–74, particularly in eastern Germany.

These findings identify older adults as a priority for liver disease surveillance and care and show that mortality registration needs better aetiological information. Further age–period–cohort analyses are needed to determine whether the observed patterns reflect the ageing of high-risk birth cohorts.

## Supporting information

STROBE checklist

## Data Availability

All data are publicly available from the WHO Mortality Database, Eurostat, the GBE-Bund health reporting system of the Federal Statistical Office and the IHME Global Health Data Exchange. The analysis scripts, query parameters and data log are provided at github.com/m-kumpf/liver-mortality-germany and archived at https://doi.org/10.5281/zenodo.24260562.

## Contributors

LA, KB and MK conceived the study. MK acquired and analysed the data, produced the figures and wrote the original draft of the manuscript. MK, LA, and KB accessed and verified the underlying data and had final responsibility for the decision to submit. MM, LA, and KB provided resources. The study was supervised by LA and KB. All authors had full access to the data, added important intellectual content by reviewing and editing the final manuscript, and approved the final version.

## Declaration of generative AI and AI-assisted technologies

During the preparation of this work the authors used Claude and Claude Code (Anthropic, Fable 5) to retrieve Global Burden of Disease Study 2023 estimates through the IHME data connector, to reformat analysis code and to edit the language of the manuscript. Retrieved estimates were checked against the documented query parameters, and all code was run and verified locally by the authors. The authors reviewed and edited the output and take full responsibility for the content of the published article.

## References

1 Karlsen TH, Sheron N, Zelber-Sagi S, et al. The EASL–Lancet Liver Commission: protecting the next generation of Europeans against liver disease complications and premature mortality. The Lancet 2022; 399: 61–116.

2 Ascione A, Fontanella L, Imparato M, Rinaldi L, De Luca M. Mortality from cirrhosis and hepatocellular carcinoma in Western Europe over the last 40 years. Liver International 2017; 37: 1193–201.

3 Naghavi M, Kyu HH, Bhoomadevi A, et al. Global burden of 292 causes of death in 204 countries and territories and 660 subnational locations, 1990–2023: a systematic analysis for the Global Burden of Disease Study 2023. The Lancet 2025; 406: 1811–72.

4 Kraus L, Østhus S, Amundsen EJ, et al. Changes in mortality due to major alcohol-related diseases in four Nordic countries, France and Germany between 1980 and 2009: a comparative age–period–cohort analysis. Addiction 2015; 110: 1443–52.

5 Piontek D, Kraus L. Trends in alcohol-related mortality in East and West Germany, 1980–2014: age, period and cohort variations. Addiction 2018; 113: 836–44.

6 Sepanlou SG, Safiri S, Bisignano C, et al. The global, regional, and national burden of cirrhosis by cause in 195 countries and territories, 1990–2017: a systematic analysis for the Global Burden of Disease Study 2017. The Lancet Gastroenterology & Hepatology 2020; 5: 245–66.

7 Delacôte C, Lenne X, Louvet A, et al. Trends in mortality and years of life lost among patients with cirrhosis and hepatocellular carcinoma in France, 2011–2023: a nationwide retrospective population-based study. The Lancet Regional Health – Europe 2026; 66. DOI:10.1016/j.lanepe.2026.101685.

8 Stolpe S, Kowall B. Todesursachenstatistik – wie Fehlinterpretationen von Mortalitätsdaten vermieden werden. Bundesgesundheitsblatt Gesundheitsforschung Gesundheitsschutz 2025; 68: 167–75.

9 Johnson SC, Cunningham M, Dippenaar IN, et al. Public health utility of cause of death data: applying empirical algorithms to improve data quality. BMC Med Inform Decis Mak 2021; 21: 175.

10 World Health Organization. WHO Mortality Database. 2026; published online Feb 23. https://www.who.int/data/data-collection-tools/who-mortality-database (xaccessed Oct 26, 2026).

11 Eurostat. Causes of death - standardised death rate by NUTS 2 region of residence. Eurostat. 2026; published online June 8. DOI:10.2908/hlth_cd_asdr2.

12 Eurostat. Population on 1 January by age and sex. Eurostat. 2026; published online Sept 25. DOI:10.2908/demo_pjan.

13 Statistisches Bundesamt. Informationssystem der Gesundheitsberichterstattung des Bundes (IS-GBE). Gesund-heitsberichterstattung des Bundes. https://www.gbe-bund.de/gbe/ (accessed Oct 27, 2026).

14 Global Burden of Disease Collaborative Network. Global Burden of Disease Study 2023 (GBD 2023). Seattle, WA, USA: Institute for Health Metrics and Evaluation, 2025 https://vizhub.healthdata.org/gbd-results (accessed Oct 27, 2026).

15 Sieber E, Héon V, Willich SN. Alkoholmissbrauch und -abhängigkeit in der ehemaligen DDR und in den neuen Bundesländern. Soz Präventivmed 1998; 43: 90–9.

16 Statistische Ämter des Bundes und der Länder. Adipositasquote. Gemeinsames Statistikportal. https://www.statistikportal.de/de/nachhaltigkeit/ergebnisse/ziel-3-gesundheit-und-wohlergehen/adipositasquote (accessed Nov 6, 2026).

17 Mensink GBM, Beitz R. Food and nutrient intake in East and West Germany, 8 years after the reunification—The German Nutrition Survey 1998. Eur J Clin Nutr 2004; 58: 1000–10.

18 Robert-Koch-Institut. Daten und Fakten: Ergebnisse der Studie ‘Gesundheit in Deutschland aktuell 2012’. Robert Koch-Institut, 2014 DOI:10.25646/3170.

19 Lange C, Manz K, Kuntz B. Alcohol consumption among adults in Germany: risky drinking levels. J Health Monit 2017; 2: 64–70.

20 Grigoriev P, Pechholdová M, Mühlichen M, Scholz RD, Klüsener S. 30 Jahre Deutsche Einheit: Errungenschaften und verbliebene Unterschiede in der Mortalitätsentwicklung nach Alter und Todesursachen. Bundesgesundheitsblatt Gesundheitsforschung Gesundheitsschutz 2021; 64: 481–90.

21 Kilian C, Rehm J, Shield K, Manthey J. Changes in Alcohol-Specific Mortality During the COVID-19 Pandemic in 14 European Countries. Sucht 2023; 69: 285–93.

22 Gu W, Hortlik H, Erasmus H-P, et al. Trends and the course of liver cirrhosis and its complications in Germany: Nationwide population-based study (2005 to 2018). Lancet Reg Health Eur 2021; 12: 100240.

23 Bengtsson B, Askling J, Ludvigsson JF, Hagström H. Validity of administrative codes associated with cirrhosis in Sweden. Scandinavian Journal of Gastroenterology 2020; 55: 1205–10.

24 Ratib S, West J, Fleming KM. Liver cirrhosis in England—an observational study: are we measuring its burden occurrence correctly? BMJ Open 2017; 7: e013752.

25 Thomadakis C, Gountas I, Duffell E, et al. Prevalence of chronic HCV infection in EU/EEA countries in 2019 using multiparameter evidence synthesis. The Lancet Regional Health – Europe 2024; 36. DOI:10.1016/j.lanepe.2023.100792.

26 Russo FP, Zanetto A, Salmaso L, et al. Decline in Complications and Mortality in Chronic Liver Disease and Cirrhosis: A Population-Based Cohort Study From Northeastern Italy. United European Gastroenterology Journal 2025; 13: 1469–79.

27 Burzyńska M, Pikala M. Changing Patterns of Digestive Disease Mortality in Older Adults in Poland, 2000–2022: Age- and Sex-Specific Trends in Liver Disease Mortality. Journal of Clinical Medicine 2026; 15: 7001.

28 Mokdad AA, Lopez AD, Shahraz S, et al. Liver cirrhosis mortality in 187 countries between 1980 and 2010: a systematic analysis. BMC Med 2014; 12: 145.

29 Tacke F, Kim Y, Haas JS, et al. Understanding the economic and healthcare burden of metabolic dysfunction-associated steatohepatitis: a real-world claims data analysis from Germany. J Comp Eff Res 2026; 15: e260092.

30 Manthey J, Kilian C, Schäfer I, Wirth M, Schulte B. Changes in the alcohol-specific disease burden during the COVID-19 pandemic in Germany: interrupted time series analyses. Eur J Public Health 2023; 33: 645–52.

