## appendix for "The plateau in liver disease mortality in Germany: contrasting trends by age, region and ICD-10 code, 2000–2024"

### Contents

#### Supplementary methods

S1 Data sources and query parameters (table S1)
S2 ICD-10 code groups (table S2)
S3 Age harmonisation and within-band standardisation
S4 Validation of the standardised rates (table S3)
S5 Analysis pipeline and data log (table S4)
S6 Exposure and survey data

#### Supplementary results

Table S5 Chronic liver disease mortality by country, Eurostat and GBD 2023
Table S6 Liver disease mortality in East and West Germany by sex and age band
Table S7 Liver disease mortality at 65–74 years by Land
Table S8 Hospital stays by main diagnosis, East and West Germany
Table S9 Self-reported drinking in eastern and western Germany by sex and age band, GEDA surveys
Figure S1 Age-specific mortality from alcoholic liver disease and from fibrosis and cirrhosis, women
Figure S2 GBD 2023 estimates against registry rates by age band
Figure S3 Alcohol and obesity exposure in eastern and western Germany
Figure S4 Self-reported drinking pattern in eastern and western Germany by age, GEDA surveys
Figure S5 Alcohol-specific mortality (K70, F10) in eastern and western Germany by sex and age band

### Supplementary methods

#### S1 Data sources and query parameters

Table S1 lists each raw input with the query or download used, the version of the source at retrieval and the content retained. Each row corresponds to an entry in the data log that accompanies the analysis scripts.

**Table S1.** Data sources, queries and versions

| Source | Table or query | Content retained | Version |
|---|---|---|---|
| WHO Mortality Database | Bulk download (ICD-10 parts 1–6, supplementary file morticd10_add, population file, country codes, availability notes) | Deaths by single ICD-10 code, sex and age group from 2000, retained for 16 European countries (K70–K77, B15–B19, C22, F10, I85, I98 and all causes). Germany reports four-character codes for 2000–2022, with age cells of fewer than three deaths suppressed from 2021 and complete all-ages totals. UK deaths 2001–2024 with Eurostat denominators to 2018 and WHO denominators for 2019, 2020, 2023 and 2024. | Release of February 22–23, 2026 |
| Eurostat | hlth_cd_asdr2 (unit RT, age TOTAL, icd10 K70_K73_K74, K72_K75, C22, B15_B19_B942, B180_B182, F10, sex T, M, F), hlth_cd_aro (deaths, residents, 33 age groups), hlth_cd_acdr2 (crude rates), demo_pjan (population on January 1 by single year of age, 2010–2025) | Published age-standardised death rates (ESP2013), 36 countries, 2011–2023 (UK to 2018), deaths by age group and population denominators. | Dataset updated June 8, 2026 |
| Federal Statistical Office, GBE-Bund, table 'Sterbefälle, Sterbeziffer (altersstandardisiert), häufigste Todesursachen (ab 1998)' (indicator 517) | One sheet per year × region × sex and age group (all ages, under 65, 65 and over), 1,539 sheets retrieved by script | Deaths and ESP2013 rates for the 100 most frequent three-character causes, 16 Länder, Germany, former federal territory with East Berlin and new Länder without East Berlin, 1998–2024. K70, K74 and C22 in every sheet, F10 in at least 90%. | Todesursachenstatistik, retrieved October 9, 2026, including 2024 |
| GBE-Bund, table 'Sterbefälle, Sterbeziffern (ab 1998), Gliederungsmerkmale Jahre, Region, Alter, Geschlecht, Nationalität, ICD-10, Art der Standardisierung' (indicator 6) | 1,539 sheets (27 years × 19 regions × 3 sexes) | Deaths and crude rates by ICD-10 block and 21 age groups. K70–K77 is the finest liver code. Cells with fewer than three deaths suppressed | Todesursachenstatistik, retrieved October 11–13, 2026 |
| GBE-Bund, 'Diagnosedaten der Krankenhäuser nach Wohnsitz (ICD10-4-Steller, ab 2000)' (indicator 703) | 1,368 sheets (24 years × 19 regions × 3 sexes), K00–K93 opened to four-character codes | Inpatient stays by four-character main diagnosis (K70.0 to K77.8) and residence, 42 age groups, 2000–2023. Stays, not persons. Full inpatient episodes including Stundenfälle. No in-hospital deaths in this table. | Krankenhausstatistik (Diagnosedaten), retrieved October 11–13, 2026 |

| Source | Table or query | Content retained | Version |
|---|---|---|---|
| GBE-Bund, 'Diagnosedaten der Krankenhäuser nach Wohnsitz (ICD-10-3-Steller, ab 2000)' (indicator 594) | 1,368 sheets, hierarchy opened for K70–K77, C15–C26, B15–B19, F10–F19 | Stays, bed days, mean length of stay and in-hospital deaths by three-character main diagnosis, 42 age groups, 2000–2023 | Krankenhausstatistik, retrieved October 12–14, 2026 |
| GENESIS-Online, tables 12411-0041 and 12411-0042 | REST API tablefile | Mid-year population of Germany by single year of age and sex (2000–2025) and by Land and sex | Retrieved October 9, 2026 |
| GBD 2023 (IHME) through the IHME data connector | get_multi_location_trend, cause 521 (cirrhosis and other chronic liver diseases), deaths, 2010–2023, 29 countries and regions by sex with 95% UI, and by five-year age group (45–49 to 95 and over) for nine countries, 2000–2023. get_ cause_burden by age for causes 521, 522 (alcohol), 523 (hepatitis B), 524 (hepatitis C), 971 (non-alcoholic fatty liver disease) and 525 (other), 2010, 2019 and 2023. get_ percent_change 2010–2023, 16 locations | Age-standardised and age-specific rates with 95% uncertainty intervals, published percent change. | GBD 2023, retrieved October 8–15, 2026 |
| Sieber E, Héon V, Willich SN. Soz Präventivmed 1998; 43: 90–99 | Tables 1 and 2 (statistical yearbooks of the GDR and the FRG), hand transcription checked against the article PDF and validated by download_exposures.py | Pure alcohol in litres per inhabitant, GDR 1955–1989 and FRG 1960–1989, GDR beer, wine and spirits in litres of beverage, spirits share of pure alcohol in 1988 from the text. The two states calculated consumption with different methods. | Article of 1998, transcribed October 29, 2026 |
| Statistische Ämter des Bundes und der Länder, statistikportal.de, SDG indicator 3 'Adipositasquote' | HTML table parsed by script, SDG Tabellenband (XLSX) archived | Share of adults (18 years and over) with a body-mass index above 30 $kg/m^2$ (Mikrozensus), 16 Länder and Germany, 1999, 2005, 2013, 2017, 2021 and 2025 (first results). From survey year 2020 only partly comparable. No sex or age split. | Page retrieved November 4, 2026 |
| WHO Regional Office for Europe, Health for All database (HFA-DB), API v5 | Measures HFA_426 (total), HFA_427 (spirits), HFA_428 (wine), HFA_429 (beer), filter COUNTRY:DEU | Recorded pure alcohol per person aged 15 years and over, Germany 1961–2019 (Federal Republic before 1990), used to cross-check the Sieber series. | Data updated October 23, 2026, retrieved November 4, 2026 |
| GBE-Bund, tables 192 'Alkoholkonsum (2009–2012)', 193 'Rauschtrinken (2009–2012)', 26612 'Alkoholkonsum (2014/2015)' and 26613 'Rauschtrinken (2014/2015)' | 120 sheets (survey year × age band × sex, education all groups) retrieved by script | GEDA 2009, 2010 and 2012 (telephone): risky drinking (AUDIT-C), heavy episodic drinking and never drinkers for Germany, western Germany, eastern Germany including Berlin and seven sub-regions. GEDA 2014/2015-EHIS (self-administered): risky intake (EHIS threshold) and heavy episodic drinking, 16 Länder. Age bands 18–29, 30–44, 45–64, 65 and over, all ages. | GEDA (Robert Koch Institute), retrieved November 4, 2026 |

| Source | Table or query | Content retained | Version |
|---|---|---|---|
| Robert Koch Institute reports and fact sheets | GEDA fact sheets 2009, 2010 and 2012 (RKI 2011, 2012, 2014). Lange C, Manz K, Kuntz B. J Health Monit 2017; 2(2): 64–70 and 71–77. Mensink GBM, Beitz R. Eur J Clin Nutr 2004; 58: 1000–1010 | Regional statements on risky and heavy episodic drinking (GEDA fact sheets), BGS98 adjusted regional means of alcohol intake by sex (transcribed). | Publications 2002–2017, retrieved October 29 to November 4, 2026 |
| GBD 2023 (IHME) through the IHME data connector, exposure calls | get_risk_attributable_burden (risk High alcohol use, cause 521, measure deaths, by age, Germany, 2010 and 2023); get_sev (High alcohol use, High body-mass index, age-standardised, Germany, 1990–2023) | Population attributable fraction of cirrhosis deaths with 95% UI by five-year age group and sex; summary exposure values with 95% UI | GBD 2023, retrieved October 29, 2026 |

UI = uncertainty interval. Retrieval scripts and parameters are listed in section S5.

### S2 ICD-10 code groups

**Table S2.** ICD-10 code groups and GBD cause definitions

| Code | Label | Sources in which it is used | Note |
|---|---|---|---|
| K70 | Alcoholic liver disease | WHO, Eurostat, GBE-Bund TOP-100 | Three-character code |
| K70.3 | Alcoholic cirrhosis of liver | WHO (Germany), hospital four-character table | |
| K74 | Fibrosis and cirrhosis of liver | WHO, GBE-Bund TOP-100 | |
| K74.6 | Other and unspecified cirrhosis of liver | WHO (Germany), hospital four-character table | 98% of K74 deaths in Germany |
| K76 | Other diseases of liver | WHO | |
| K76.0 | Fatty (change of) liver, not elsewhere classified | WHO (Germany), hospital four-character table | Closest routine code for metabolic fatty liver disease |
| K70, K73, K74 | Chronic liver disease (Eurostat shortlist) | Eurostat hlth_cd_asdr2, own standardisation of WHO counts | Used for the validation against Eurostat |
| K70–K76 | Alcoholic liver disease to other diseases of liver | WHO | Closest registry equivalent of the GBD cause cirrhosis and other chronic liver diseases |
| K70–K77 | Diseases of liver | GBE-Bund block table, hospital three-character table | Finest liver code available by five-year age group and Land |
| C22 | Malignant neoplasm of liver and intrahepatic bile ducts | WHO, Eurostat, GBE-Bund, hospital | |
| B18 | Chronic viral hepatitis | WHO, Eurostat, hospital | |
| F10 | Mental and behavioural disorders due to use of alcohol | WHO, Eurostat, GBE-Bund, hospital | |
| GBD cause 521 | Cirrhosis and other chronic liver diseases | GBD 2023 | Total estimated by CODEm. Sub-causes 522 alcohol, 523 hepatitis B, 524 hepatitis C, 971 non-alcoholic fatty liver disease, 525 other causes, assigned by modelled proportions |

ICD-10 labels follow the WHO 2019 version. German registry data use the German modification (ICD-10-GM) for hospital diagnoses and the WHO version for causes of death.

### S3 Age harmonisation and within-band standardisation

The WHO Mortality Database reports deaths in up to 26 age columns whose meaning depends on the age format of the row (formats 00 to 03 were used). Single-year columns under five years were summed to under 1 year and 1–4 years, and the open-ended top group was taken as the last non-empty column of the all-cause row of the same country, year and sex. That top group (85, 90 or 95 years and over depending on country and year) was applied to every cause of the same country, year and sex. Deaths of unknown age were redistributed in proportion to the deaths of known age. From 2021 the German file suppresses age-specific cells with fewer than three deaths but reports the all-ages total of every row. The suppressed counts were imputed by scaling the reported age-specific counts of each row (ICD-10 code, sex and year) to the reported total, which allocates the missing deaths across age groups in proportion to the reported counts. Without this step the 2021 and 2022 counts would be 1.3% low for K70 to K76 (220 and 234 deaths), about a quarter low for K76.0 and below 0.6% low for K70, K74 and K74.6. Rows in which every age cell is suppressed (75 and 51 deaths, four-character codes with at most 11 deaths each) and rows whose total is suppressed (at most 18 and 28 deaths) remain uncounted. Allocating the missing deaths to the suppressed cells alone instead changes the 2022 band rates for K70 and K74 at 45–64 and 65–74 years by at most 0.2 per 100 000 and their 2010–2022 changes by at most 1.0 percentage point.

Denominators are Eurostat mid-year populations by single year of age (mean of the stocks on January 1 of the year and of the following year), aggregated to the 21 age groups. Rates were directly standardised to the European Standard Population 2013 (ESP2013, 21 groups) and, for the comparison with GBD, to the WHO world standard population. Where the top group of a row is wider than 95 years and over, the standard population weights above the top group were merged into it.

Age-band rates for 45–64, 65–74 and 75 years and over are ESP2013-weighted means of the five-year crude rates within the band, with the weights 7000, 7000, 6500 and 6000 (45–49 to 60–64), 5500 and 5000 (65–69 and 70–74) and 4000, 2500, 1500, 800 and 200 (75–79 to 95 years and over). The German band rates from the WHO file match the GBE-Bund age-band sheets (2010, K70, under 65 years 8.3 against 8.3 per 100 000, 65 years and over 18.6 against 18.5). GBD five-year rates and GBE-Bund crude age-specific rates were combined to the same bands with the same weights. At 75 years and over the top group differs by source (85 years and over for the German WHO rows from 2017, 90 for GBE-Bund, 95 for GBD). Three-year means for the Länder are arithmetic means of the annual band rates and differ from rates pooled over the three years by less than 1% in every Land and sex at 65–74 years.

### S4 Validation of the standardised rates

**Table S3.** Own ESP2013 age-standardised death rates for K70, K73 and K74 combined against the rates published by Eurostat, both sexes, by country

| Country | Years compared | n | Median \|difference\| (%) | Maximum \|difference\| (%) | Mean signed difference (%) |
|---|---|---|---|---|---|
| Austria | 2011–2023 | 13 | 0.35 | 0.72 | −0.39 |
| Belgium | 2011–2022 | 12 | 0.69 | 1.06 | −0.64 |
| Bulgaria | 2011–2023 | 13 | 0.48 | 1.44 | +0.40 |
| Croatia | 2011–2023 | 12 | 0.31 | 5.71 | +0.59 |
| Cyprus | 2011–2023 | 13 | 1.75 | 4.21 | −0.19 |
| Czechia | 2011–2023 | 13 | 0.15 | 0.33 | −0.17 |
| Denmark | 2011–2022 | 12 | 0.73 | 3.86 | −1.02 |
| Estonia | 2011–2023 | 13 | 0.22 | 2.70 | −0.54 |
| Finland | 2011–2023 | 13 | 0.38 | 1.01 | +0.34 |
| France | 2011–2023 | 13 | 2.57 | 3.88 | −2.57 |
| Germany | 2011–2022 | 12 | 0.22 | 0.40 | −0.22 |
| Greece | 2014–2022 | 9 | 1.55 | 2.34 | +1.45 |
| Hungary | 2011–2023 | 13 | 0.70 | 1.53 | +0.71 |
| Iceland | 2011–2023 | 13 | 4.28 | 29.66 | −2.75 |
| Ireland | 2011–2022 | 12 | 0.47 | 2.22 | −0.32 |
| Italy | 2011–2023 | 13 | 1.26 | 2.24 | +1.22 |
| Latvia | 2011–2023 | 13 | 0.28 | 1.59 | −0.36 |
| Lithuania | 2011–2023 | 13 | 0.87 | 1.70 | −0.46 |
| Luxembourg | 2011–2023 | 13 | 1.96 | 10.52 | −3.45 |
| Malta | 2011–2022 | 12 | 0.77 | 5.25 | +0.61 |
| Netherlands | 2011–2023 | 13 | 0.65 | 1.57 | −0.82 |
| Norway | 2011–2016 | 6 | 1.96 | 2.47 | −1.72 |
| Poland | 2011–2023 | 13 | 0.38 | 1.12 | −0.23 |
| Portugal | 2011–2022 | 10 | 0.34 | 1.39 | −0.17 |
| Romania | 2011–2023 | 13 | 0.12 | 0.88 | −0.01 |
| Slovakia | 2012–2023 | 11 | 1.30 | 2.10 | −1.32 |
| Slovenia | 2011–2023 | 13 | 1.22 | 2.72 | −1.23 |
| Spain | 2011–2023 | 13 | 0.89 | 1.27 | +0.84 |
| Sweden | 2011–2023 | 13 | 0.37 | 0.98 | −0.22 |
| Switzerland | 2011–2023 | 13 | 0.56 | 1.35 | −0.68 |
| United Kingdom | 2011–2018 | 8 | 0.59 | 0.95 | −0.03 |

Difference = (own rate − Eurostat rate) / Eurostat rate × 100, from WHO Mortality Database counts and Eurostat mid-year populations. The median absolute difference is below 1% for 22 of 31 countries. Larger differences for France (territory definition), Greece, and Iceland, Luxembourg, Malta and Cyprus (small numbers) reflect population and territory definitions rather than the method. German death counts in the GBE-Bund tables are identical to the WHO counts up to 2020 and, after the rescaling of section S3, within 0.3% for K70 and K74 in 2021 and 2022.

### S5 Analysis pipeline and data log

All steps from download to tables are numbered Python scripts in the folder 20_analysis of the project. Each script reads only raw files or the output of an earlier step and writes only to the derived-data folder. The data log (00_meta/data_log.md) records for every file the date of retrieval, the query or script, the version of the source and validation notes. Scripts, data log, derived tables and figure files are available at github.com/m-kumpf/liver-mortality-germany (version 1.0.0, archived at https://doi.org/10.5281/zenodo.24260562).

**Table S4.** Analysis scripts in the order of execution

| Step | Script | Output |
|---|---|---|
| 1 | download_eurostat.py | Eurostat API pulls (hlth_cd_asdr2, hlth_cd_aro, hlth_cd_acdr2, demo_pjan) |
| 2 | extract_who_mdb.py | Liver, viral hepatitis, liver cancer, alcohol use disorder and all-cause rows from the WHO Mortality Database, harmonised age groups |
| 3 | standardise_who_mdb.py | Rescaling of suppressed German age cells, standardisation to the ESP2013 and the WHO world standard, validation against Eurostat (table S3), coding indicators (K74 share, K76.0 share) |
| 4 | download_destatis.py | GENESIS population tables |
| 5 | download_gbe_bund.py | GBE-Bund TOP-100 cause-of-death table by Land, sex and age group |
| 6 | age_specific_comparison.py | Age-band rates for 16 countries, 2000–2023 (Germany to 2022) |
| 7 | age_band_detail.py | Bands 45–64, 65–74 and 75 and over for K70, K74, K74.6 and K70 plus K74 by sex, nine countries (table 1, figures 2 and S1) |
| 8 | gbd_vs_registry_age.py | GBD five-year rates combined to the same bands and compared with the registry (figure S2) |
| 9 | east_west_age_bands.py | K70–K77 band rates for East and West Germany (figure 3A, B, table S6) |
| 10 | hospital_age_bands.py | Hospital stays by four-character main diagnosis and band, East and West (figure 4A, table S8) |
| 11 | hospital3_bands.py | Three-character hospital table with in-hospital deaths (figure 4B, C, table S8) |
| 12 | laender_age_bands.py | Band rates by Land, three-year means, coding shares (figure 3C, D, table S7) |
| 14 | make_tables.py | Manuscript tables as printed |
| 15 | gbd_standard_check.py | GBD age-specific rates re-standardised to the WHO world standard and the GBD-to-registry ratio on one standard, 26 country-years (table S5 footnote) |
| 16 | download_exposures.py | Mikrozensus obesity shares by Land, WHO Europe HFA-DB alcohol series, validation of the Sieber transcription (figure S3) |
| 17 | exposures_east_west.py | Annual GDR and FRG series with interpolation, cumulative consumption by birth cohort, East and West obesity means, GBD attributable fractions and summary exposure values (figure S3) |
| 18 | download_gbe_geda.py | GBE-Bund GEDA tables 192, 193, 26612 and 26613 (120 sheets) and RKI report statements |
| 19 | geda_east_west.py | East and West estimates by indicator, sex and age band, mean of the 2009, 2010 and 2012 waves, population-weighted Länder means for 2014/2015 (figure S4) |
| 20 | alcohol_mortality_east_west.py | K70 and F10 mortality in East and West Germany by sex and age band, 1998–2024, with recovery of the women's F10 cells at 65 and over (figure S5) |

Shared helpers for age harmonisation, the rescaling of suppressed cells, standard populations and Eurostat denominators are in whomdb_common.py. GBE-Bund tables were retrieved with download_gbe_bund.py, download_gbe_full.py and download_gbe_geda.py. The final run used Python 3.13, pandas 2.3.3, numpy 2.2.6 and openpyxl.

#### S6 Exposure and survey data

Recorded consumption of pure alcohol per inhabitant in the German Democratic Republic (GDR, 1955–1989) and the Federal Republic of Germany (FRG, 1960–1989) was transcribed from tables 1 and 2 of Sieber and colleagues, who compiled the statistical yearbooks of both states, and checked against the article. The two states calculated consumption with different methods. The FRG series agrees with the WHO Europe Health for All database (ratio 0.70 to 0.81 in 1970–1989, consistent with the share of the population aged 15 years and over). Cumulative consumption per inhabitant to 1989 was summed for the birth cohorts 1940–44 to 1955–59 from the year in which the mid-cohort birth year reached age 15, starting no earlier than 1960, with linear interpolation between reported years, and was 26–41% higher in the FRG. Per-capita sales do not measure individual intake (figure S3A, B).

Adult obesity prevalence (body-mass index above 30 kg/m$^2$, ages 18 years and over) by Land is the Mikrozensus indicator published by the Statistische Ämter des Bundes und der Länder for 1999, 2005, 2013, 2017, 2021 and 2025. Results from survey year 2020 onwards are only partly comparable with earlier years, and the 2025 values are first results. Eastern and western values are unweighted means of the five eastern and the ten western Länder, Berlin excluded. Population-weighted means differ from them by at most 0.5 percentage points (figure S3C).

The GBD 2023 population attributable fraction of cirrhosis deaths for high alcohol use by five-year age group and sex, and the age-standardised summary exposure values for high alcohol use and high body-mass index, were retrieved for Germany through the IHME data connector (table S1). The attributable fraction is derived from the GBD aetiology proportions, and its decline of one to two points between 2010 and 2023 lies within the uncertainty interval (figure S3D). GBD 2023 does not model high body-mass index as a risk factor for cirrhosis deaths.

Self-reported drinking by region was taken from the GEDA surveys of the Robert Koch Institute through GBE-Bund (table S1). GEDA 2009, 2010 and 2012 were telephone interviews with 19 000 to 22 000 respondents per wave. Risky drinking is an AUDIT-C score of at least 5 in men and 4 in women, and heavy episodic drinking six or more alcoholic drinks on one occasion at least weekly. These waves are published for Germany, western Germany and eastern Germany including Berlin, whereas the mortality definition assigns East Berlin to the West, and were summarised as the mean of the three published estimates. GEDA 2014/2015-EHIS was a self-administered questionnaire with 24 016 respondents. Risky intake is more than 20 g (men) or 10 g (women) of pure alcohol per day among people who drink at least weekly, as a share of all respondents, and heavy episodic drinking six or more drinks on one occasion at least weekly in the past 12 months. This survey is published by Land, and eastern and western values are population-weighted means of the five eastern and the ten western Länder, Berlin excluded. The two survey generations differ in instrument, answer categories, reference period and mode, so regions are compared within a survey generation only.

GBE-Bund and the RKI reports publish no regional confidence intervals, so the regional estimates and their differences are reported without intervals. The RKI reports describe risky drinking among men in the southern eastern region as significantly above the federal average in 2010 and no regional differences in weekly heavy episodic drinking in 2012.

The Bundes-Gesundheitssurvey 1998 (BGS98, 7 124 participants aged 18–79 years) measured alcohol intake in a dietary history interview of the nutrition sub-sample. Mensink and Beitz reported regional means adjusted for age, socioeconomic status, smoking, physical activity, season and energy intake of 19.2 g per day (95% CI 17.2–21.3) in eastern and 16.8 g per day (15.7–17.9) in western men and of 6.3 and 6.4 g per day in women. BGS98 and the two GEDA generations use different instruments, so no trend is derived across them.

Mortality from alcoholic liver disease (K70) and from mental and behavioural disorders due to alcohol (F10) in eastern and western Germany was taken from the GBE-Bund table of the 100 most frequent three-character causes of death (indicator 517, table S1) for 1998–2024, which publishes age-standardised rates (ESP2013) for all ages and within the bands under 65 and 65 years and over. F10 for women aged 65 years and over is absent from the list of 100 causes in most years (25 of 27 in the East). For these cells the number of deaths was recovered exactly as all ages minus under 65 and the rate estimated by distributing the deaths over the five-year age groups in proportion to the F10–F19 block of the full table. In the 436 published cells on which this allocation could be tested, the median relative difference was +0.1% and 90% of estimates were within 8.3% of the published rate. The share of F10 among K70 and F10 deaths combined is higher in the West than in the East (men 46% against 34% in 2024, women 39% against 23%), so the East to West ratio for K70 alone overstates the alcohol-specific excess (men aged 65 and over in 2024, 1.78 for K70 and 1.37 for K70 and F10 combined; figure S5).

### Supplementary results

**Table S5.** Age-standardised mortality from chronic liver disease by country: Eurostat registry series and GBD 2023 estimates

| Country | Eurostat 2011 | Eurostat 2019 | Eurostat 2023 | Eurostat change | GBD 2010 | GBD 2019 | GBD 2023 | GBD change (95% UI) | GBD / registry |
|---|---|---|---|---|---|---|---|---|---|
| Germany | 16.6 | 15.2 | 16.0 | −4% | 12.6 | 11.7 | 12.8 | +1.9% (−6.5 to +13.2) | 1.22 |
| Austria | 17.1 | 14.3 | 14.8 | −13% | 14.2 | 10.7 | 10.5 | −25.6% (−32.7 to −16.7) | 1.16 |
| Belgium | 11.2 | 11.3 | 11.5 | +2% | 9.6 | 8.8 | 8.3 | −13.7% (−21.9 to −4.4) | 1.15 |
| Czechia | 18.5 | 19.4 | 19.5 | +6% | 14.4 | 14.4 | 14.5 | +0.3% (−8.7 to +9.5) | 1.01 |
| Denmark | 14.8 | 12.4 | 11.2 | −24% | 11.9 | 8.6 | 8.5 | −28.0% (−35.7 to −19.4) | 1.09 |
| Finland | 20.8 | 19.7 | 19.5 | −6% | 14.4 | 12.4 | 13.0 | −9.8% (−15.4 to −3.3) | 0.98 |
| France | 12.0 | 10.0 | 9.4 | −21% | 10.0 | 7.9 | 7.9 | −21.3% (−30.6 to −11.6) | 1.24 |
| Italy | 10.4 | 7.3 | 6.5 | −37% | 10.1 | 7.2 | 6.6 | −34.8% (−40.5 to −27.2) | 1.42 |
| Netherlands | 5.1 | 5.0 | 5.2 | +2% | 4.2 | 4.0 | 4.0 | −5.1% (−12.9 to +4.6) | 1.14 |
| Poland | 17.7 | 19.9 | 20.7 | +17% | 15.2 | 16.2 | 16.6 | +9.4% (+0.4 to +21.2) | 1.11 |
| Portugal | 12.6 | 10.0 | 7.9 | −37% | 12.5 | 9.1 | 8.3 | −33.8% (−41.6 to −23.8) | 1.18 |
| Spain | 11.0 | 8.2 | 7.5 | −31% | 9.6 | 6.9 | 6.5 | −32.8% (−39.7 to −25.1) | 1.21 |
| Sweden | 6.5 | 7.3 | 7.7 | +19% | 5.3 | 5.2 | 5.3 | +0.3% (−9.4 to +12.8) | 1.13 |
| Switzerland | 8.2 | 7.1 | 6.2 | −25% | 6.6 | 5.1 | 4.4 | −34.2% (−40.3 to −27.0) | 1.19 |
| United Kingdom | 12.7 | .. | 12.2 (2018) | −4% (2011–2018) | 9.8 | 9.3 | 11.5 | +17.0% (+8.4 to +27.3) | 0.95 |
| Western Europe | .. | .. | .. | .. | 10.1 | 8.4 | 8.7 | −14.0% (−19.8 to −7.2) | .. |

Eurostat columns are deaths per 100 000 from K70, K73 and K74 (underlying cause, both sexes, European Standard Population 2013). The UK series ends in 2018. GBD columns are cirrhosis and other chronic liver diseases, age-standardised to the GBD world population age standard as published, with the published change 2010–2023 and its 95% uncertainty interval. For Germany the interval includes no change and a decrease, so the +1.9% is an increase in the point estimate only. The age-band changes and the changes of the five modelled sub-causes in the main text are differences between GBD point estimates and carry no published interval. GBD / registry is the mean of the annual ratios 2015–2022 (UK 2015–2020) of the published GBD rate to the registered K70–K76 rate standardised to the WHO world standard (WHO Mortality Database). Re-standardising GBD's age-specific rates to the WHO world standard for 26 country-years (nine countries, 2010, 2019 and 2023, step 15) changes the ratio by less than 0.01 for Germany and by at most 0.07 (Italy). The within-band ratios in figure S2 use identical ESP2013 weights for GBD and registry rates. '..' not available.

**Table S6.** Mortality from diseases of the liver (K70–K77) in East and West Germany by sex and age band, 2010, 2019 and 2024

| Sex | Age (years) | East 2010 | East 2019 | East 2024 | East change | West 2010 | West 2019 | West 2024 | West change | East/West 2010 | East/West 2024 |
|---|---|---|---|---|---|---|---|---|---|---|---|
| Men | 45–64 | 68.6 | 59.9 | 55.9 | −19% | 37.4 | 30.6 | 30.0 | −20% | 1.83 | 1.86 |
| | 65–74 | 82.1 | 92.2 | 96.8 | +18% | 58.6 | 60.8 | 62.0 | +6% | 1.40 | 1.56 |
| | ≥75 | 76.7 | 71.9 | 80.3 | +5% | 62.1 | 66.7 | 67.5 | +9% | 1.24 | 1.19 |
| Women | 45–64 | 22.5 | 18.1 | 20.8 | −8% | 17.7 | 13.6 | 13.5 | −24% | 1.27 | 1.54 |
| | 65–74 | 29.7 | 31.3 | 37.9 | +28% | 26.8 | 28.7 | 30.1 | +12% | 1.11 | 1.26 |
| | ≥75 | 37.7 | 38.9 | 41.8 | +11% | 33.5 | 37.7 | 37.2 | +11% | 1.13 | 1.12 |

Deaths per 100 000 standardised to the ESP2013 within the age band (Todesursachenstatistik via GBE-Bund). East comprises the new Länder without East Berlin and West the former federal territory with East Berlin. Change 2010–2024.

**Table S7.** Mortality from diseases of the liver (K70–K77) at 65–74 years by Land, three-year means 2009–11, 2018–20 and 2022–24

| Land | Men 2009–11 | Men 2018–20 | Men 2022–24 | Men change | Men deaths/ year | Women 2009–11 | Women 2018–20 | Women 2022–24 | Women change | Women deaths/ year | K74 share, all ages |
|---|---|---|---|---|---|---|---|---|---|---|---|
| Baden-Württemberg | 76.0 | 69.7 | 70.6 | −7% | 399 | 29.1 | 30.4 | 30.5 | +5% | 191 | 0.52 |
| Bavaria | 63.6 | 66.6 | 69.6 | +10% | 466 | 28.0 | 28.5 | 31.9 | +14% | 236 | 0.52 |
| Berlin | 62.2 | 70.6 | 64.3 | +3% | 100 | 34.2 | 31.0 | 31.4 | −8% | 57 | 0.45 |
| Brandenburg (East) | 77.6 | 89.5 | 95.1 | +23% | 156 | 29.4 | 34.2 | 37.2 | +27% | 67 | 0.43 |
| Bremen | 55.0 | 61.6 | 68.4 | +24%* | 23 | 38.1 | 25.1 | 31.0 | −19%* | 12 | 0.40 |
| Hamburg | 53.2 | 46.6 | 44.2 | −17% | 32 | 32.3 | 20.1 | 26.4 | −18%* | 22 | 0.37 |
| Hesse | 57.4 | 55.9 | 56.2 | −2% | 181 | 25.7 | 25.0 | 28.5 | +11% | 101 | 0.47 |
| Lower Saxony | 53.4 | 56.4 | 60.4 | +13% | 267 | 23.0 | 28.3 | 32.2 | +40% | 155 | 0.42 |
| Mecklenburg-Western Pomerania (East) | 98.1 | 109.6 | 125.4 | +28% | 139 | 29.7 | 36.2 | 47.7 | +61% | 58 | 0.29 |
| North Rhine-Westphalia | 53.0 | 58.0 | 60.9 | +15% | 568 | 25.2 | 29.6 | 31.9 | +27% | 336 | 0.48 |
| Rhineland-Palatinate | 65.5 | 64.8 | 66.5 | +2% | 160 | 25.3 | 27.0 | 30.4 | +21% | 78 | 0.51 |
| Saarland | 72.4 | 74.8 | 73.5 | +2% | 47 | 22.3 | 42.0 | 39.9 | +79%* | 28 | 0.58 |
| Saxony (East) | 87.3 | 100.1 | 103.6 | +19% | 257 | 28.0 | 33.4 | 38.2 | +36% | 109 | 0.33 |
| Saxony-Anhalt (East) | 89.1 | 89.6 | 113.3 | +27% | 162 | 32.0 | 31.8 | 38.2 | +19% | 63 | 0.24 |
| Schleswig-Holstein | 42.4 | 54.6 | 62.1 | +47% | 100 | 21.6 | 29.0 | 34.0 | +58% | 61 | 0.36 |
| Thuringia (East) | 68.9 | 83.3 | 96.1 | +39% | 136 | 28.3 | 30.4 | 31.1 | +10% | 50 | 0.52 |

Deaths per 100 000 standardised to the ESP2013 within the band. Change between the 2009–11 and 2022–24 means. Deaths/year = mean annual deaths 2022–24. * marks fewer than 30 deaths per year, change not interpreted. K74 share = K74 deaths divided by K70 plus K74 deaths, death counts of all ages and both sexes combined, mean of 2018–20, from the GBE-Bund TOP-100 cause-of-death table. It is not restricted to 65–74 years and is not a ratio of standardised rates.

**Table S8.** Hospital stays by main diagnosis and residence, East and West Germany, both sexes, 2010, 2019 and 2023

| Main diagnosis<br>Age (years) | East 2010 | East 2019 | East 2023 | East change 2010–2019 | West 2010 | West 2019 | West 2023 | West change 2010–2019 |
|---|---|---|---|---|---|---|---|---|
| *Alcoholic cirrhosis (K70.3)* | | | | | | | | |
| 45–64 | 126.3 | 113.5 | 109.6 | −10% | 74.5 | 66.9 | 67.9 | −10% |
| 65–74 | 109.3 | 124.1 | 131.8 | +14% | 79.0 | 84.5 | 83.1 | +7% |
| ≥75 | 30.6 | 38.8 | 37.5 | +27% | 29.7 | 35.8 | 36.8 | +20% |
| *Unspecified cirrhosis (K74.6)* | | | | | | | | |
| 45–64 | 28.5 | 31.3 | 28.6 | +10% | 31.5 | 29.5 | 28.7 | −6% |
| 65–74 | 60.8 | 77.8 | 81.3 | +28% | 53.0 | 68.1 | 66.0 | +28% |
| ≥75 | 63.4 | 77.6 | 68.7 | +22% | 55.3 | 64.0 | 61.9 | +16% |
| *Fatty liver (K76.0)* | | | | | | | | |
| 45–64 | 5.3 | 5.3 | 3.7 | +1% | 3.4 | 3.3 | 2.5 | −4% |
| 65–74 | 4.5 | 3.9 | 3.0 | −15% | 3.1 | 3.3 | 2.4 | +5% |
| ≥75 | 2.4 | 2.3 | 1.1 | −4% | 2.1 | 2.2 | 1.3 | +4% |
| *Alcohol use disorders (F10)* | | | | | | | | |
| 45–64 | 741.4 | 659.4 | 546.7 | −11% | 637.4 | 552.7 | 451.3 | −13% |
| 65–74 | 168.8 | 218.4 | 208.4 | +29% | 204.5 | 213.0 | 179.0 | +4% |
| ≥75 | 40.9 | 50.7 | 49.8 | +24% | 59.1 | 62.2 | 46.4 | +5% |
| *Liver cancer (C22)* | | | | | | | | |
| 45–64 | 40.2 | 39.6 | 35.1 | −1% | 36.1 | 30.8 | 26.7 | −15% |
| 65–74 | 121.6 | 145.7 | 142.2 | +20% | 107.0 | 121.3 | 105.3 | +13% |
| ≥75 | 110.3 | 124.1 | 123.0 | +13% | 94.9 | 114.9 | 114.1 | +21% |
| *Chronic viral hepatitis (B18)* | | | | | | | | |
| 45–64 | 3.0 | 0.8 | 0.4 | −75% | 6.7 | 1.1 | 0.8 | −84% |
| 65–74 | 1.3 | 0.5 | 0.5 | −59% | 3.6 | 0.8 | 0.6 | −76% |
| ≥75 | 1.1 | 0.5 | 0.7 | −54% | 1.8 | 0.6 | 0.4 | −65% |
| *In-hospital deaths per 100 stays, K70–K77* | | | | | | | | |
| 45–64 | 10.9 | 9.1 | 9.6 | −16% | 10.4 | 8.5 | 8.5 | −19% |
| 65–74 | 13.1 | 10.4 | 12.0 | −20% | 12.4 | 9.8 | 9.7 | −21% |
| ≥75 | 17.5 | 12.2 | 12.0 | −30% | 14.9 | 12.0 | 11.9 | −20% |

Stays per 100 000 standardised to the ESP2013 within the age band (Krankenhausdiagnosestatistik via GBE-Bund). Stays are counted per admission, not per patient. Full inpatient episodes, including Stundenfälle; outpatient and partial inpatient episodes are excluded. In 2019, 88 715 stays in Germany had a main diagnosis in K70–K77, 32 547 of K70.3, 21 020 of K74.6 and 1 936 of K76.0 (all ages, both sexes). The last block gives in-hospital deaths per 100 stays for diseases of the liver (K70–K77).

**Table S9.** Self-reported drinking in eastern and western Germany by sex and age band, GEDA surveys

| Indicator and survey<br>Sex | Age (years) | Eastern Germany (%) | Western Germany (%) | Difference, percentage points |
|---|---|---|---|---|
| *Risky drinking, AUDIT-C, GEDA 2009–2012 (mean of three waves)* | | | | |
| Men | All | 36.7 | 31.9 | +4.8 |
| Men | 45–64 | 39.2 | 29.1 | +10.1 |
| Men | 65 and over | 27.1 | 29.1 | −2.0 |
| Women | All | 20.4 | 21.6 | −1.3 |
| Women | 45–64 | 21.2 | 21.4 | −0.2 |
| Women | 65 and over | 13.5 | 18.9 | −5.4 |
| *Risky intake, EHIS threshold, GEDA 2014/2015-EHIS* | | | | |
| Men | All | 20.4 | 17.5 | +2.9 |
| Men | 45–64 | 24.0 | 20.8 | +3.2 |
| Men | 65 and over | 17.2 | 18.2 | −1.0 |
| Women | All | 12.0 | 14.1 | −2.1 |
| Women | 45–64 | 14.8 | 17.5 | −2.7 |
| Women | 65 and over | 5.9 | 13.6 | −7.7 |
| *Heavy episodic drinking at least weekly, GEDA 2009–2012 (mean of three waves)* | | | | |
| Men | All | 5.4 | 5.6 | −0.2 |
| Men | 45–64 | 5.3 | 4.1 | +1.2 |
| Men | 65 and over | 3.3 | 4.7 | −1.4 |
| Women | All | 1.4 | 1.5 | −0.1 |
| Women | 45–64 | 1.6 | 1.4 | +0.2 |
| Women | 65 and over | 0.6 | 1.3 | −0.7 |
| *Heavy episodic drinking at least weekly, GEDA 2014/2015-EHIS* | | | | |
| Men | All | 12.7 | 14.3 | −1.6 |
| Men | 45–64 | 10.9 | 13.9 | −3.0 |
| Men | 65 and over | 10.1 | 13.3 | −3.3 |
| Women | All | 5.2 | 5.8 | −0.6 |
| Women | 45–64 | 5.2 | 5.6 | −0.5 |
| Women | 65 and over | 3.4 | 6.6 | −3.2 |

Eastern Germany includes Berlin in GEDA 2009–2012 (published aggregate) and excludes it in GEDA 2014/2015-EHIS (population-weighted mean of the five eastern Länder). Risky drinking: AUDIT-C score of at least 5 (men) or 4 (women). Risky intake: more than 20 g (men) or 10 g (women) of pure alcohol per day among weekly drinkers, share of all respondents. Heavy episodic drinking: six or more drinks on one occasion. Levels are not comparable between the two survey generations. Source: GBE-Bund tables 192, 193, 26612 and 26613; RKI reports GEDA 2009, 2010 and 2012; Lange, Manz and Kuntz 2017.

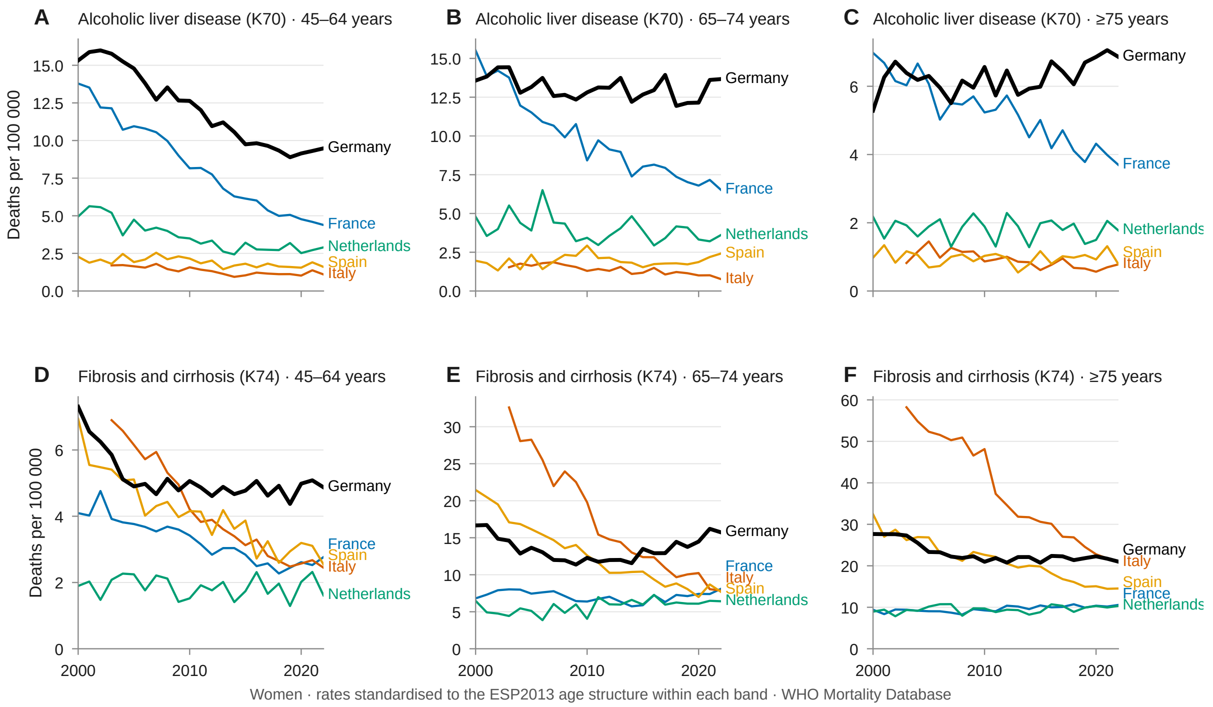


**Figure S1. Age-specific mortality from alcoholic liver disease and from fibrosis and cirrhosis, women, 2000–2022.** Deaths per 100 000 for ICD-10 K70 (A–C) and K74 (D–F) in the age bands 45–64, 65–74 and 75 years and over, standardised to the ESP2013 age structure within each band, for Germany, France, Italy, Spain and the Netherlands (WHO Mortality Database, Eurostat mid-year population). Men are shown in figure 2 of the main text.

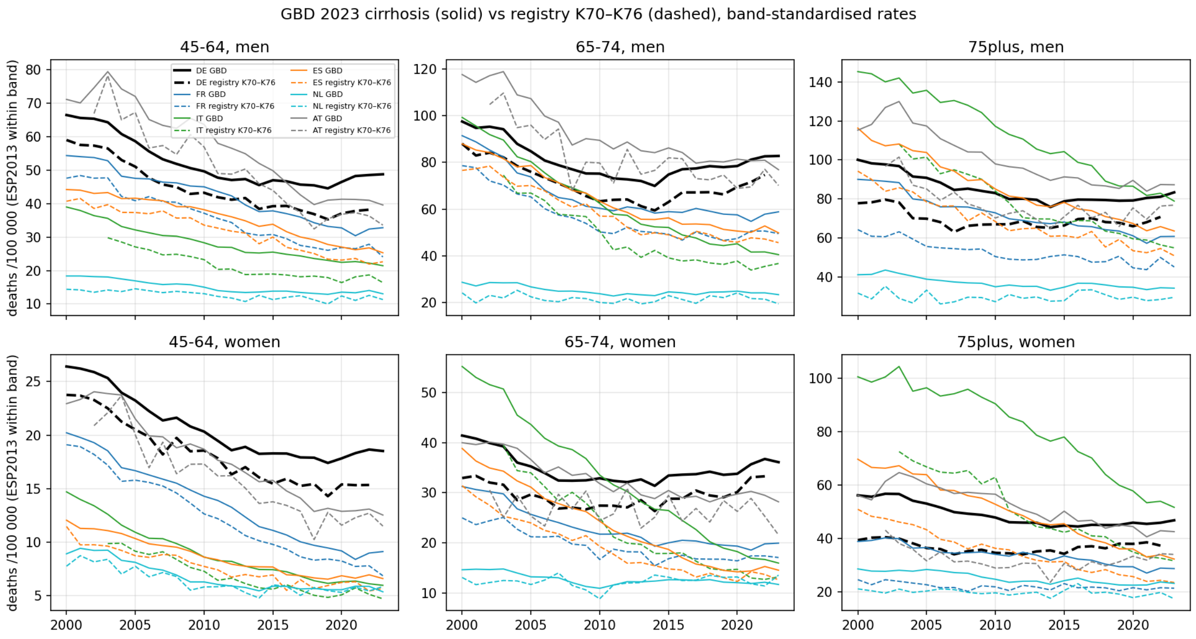


**Figure S2. GBD 2023 estimates against registry rates by age band and sex, 2000–2023 (GBD) and 2000–2022 (registry).** Deaths per 100 000 standardised to the ESP2013 within the age band for the GBD cause cirrhosis and other chronic liver diseases (solid) and the registered underlying-cause rate for K70–K76 (dashed) in Germany, France, Italy, Spain, the Netherlands and Austria. GBD five-year rates were combined to the bands with the same weights as the registry rates.

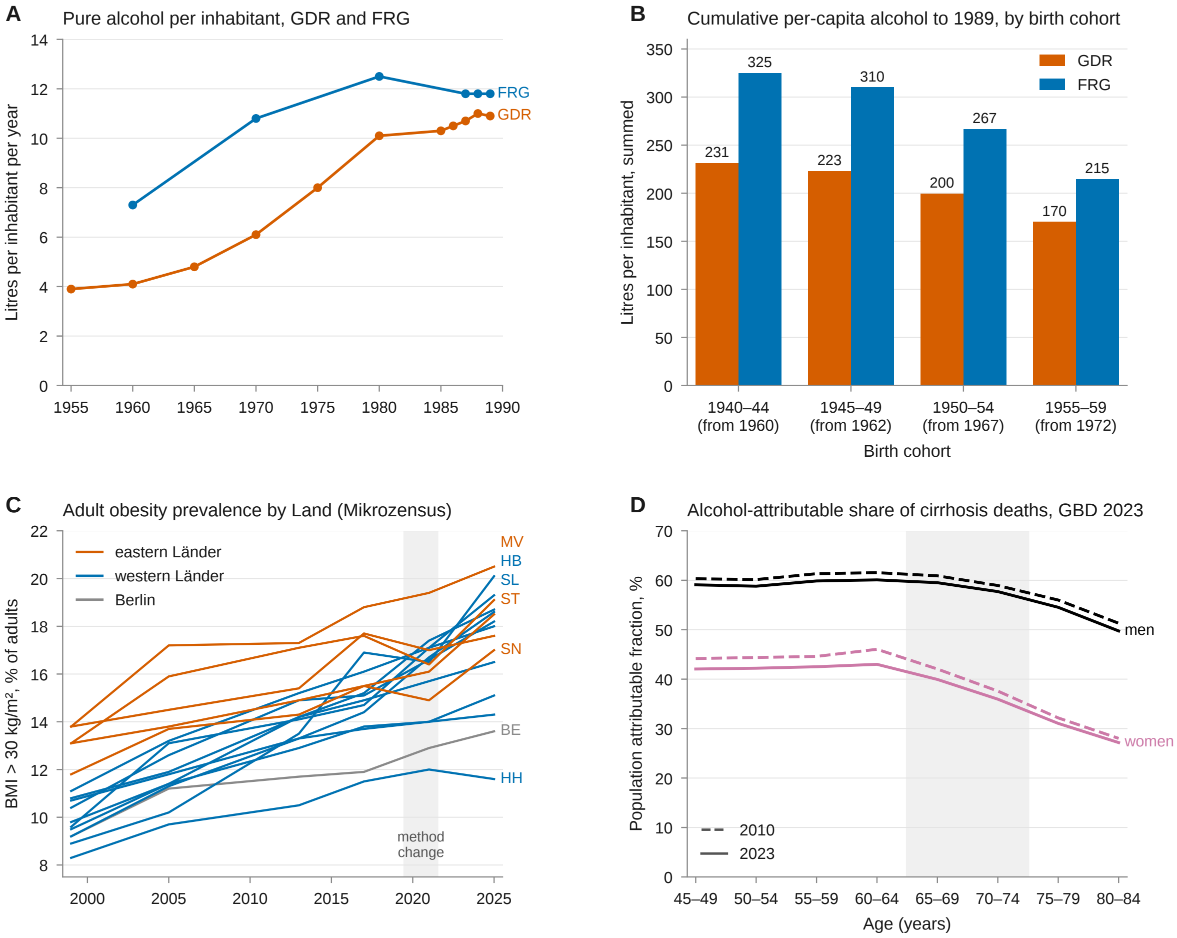


**Figure S3. Alcohol and obesity exposure in eastern and western Germany.** (A) Recorded consumption of pure alcohol in litres per inhabitant per year in the German Democratic Republic (GDR, 1955–1989) and the Federal Republic of Germany (FRG, 1960–1989), from the statistical yearbooks of both states as compiled by Sieber and colleagues; the two states calculated consumption with different methods. (B) Per-capita consumption summed from age 15 years, starting no earlier than 1960, to 1989 for the birth cohorts 1940–44 to 1955–59, with linear interpolation between reported years. (C) Share of adults (18 years and over) with a body-mass index above 30 kg/m$^2$ by Land, Mikrozensus 1999–2025; eastern Länder in vermilion, western Länder in blue, Berlin in grey. Labels mark the two highest and the lowest Länder of each group in 2025 (MV, ST, SN; HB, SL, HH) and Berlin. Results from survey year 2020 onwards are only partly comparable with earlier years (shaded), and the 2025 values are first results. (D) Share of deaths from cirrhosis and other chronic liver diseases attributed to high alcohol use by five-year age group and sex, Germany, GBD 2023 (population attributable fraction, 2010 dashed and 2023 solid; ages 65–74 shaded).

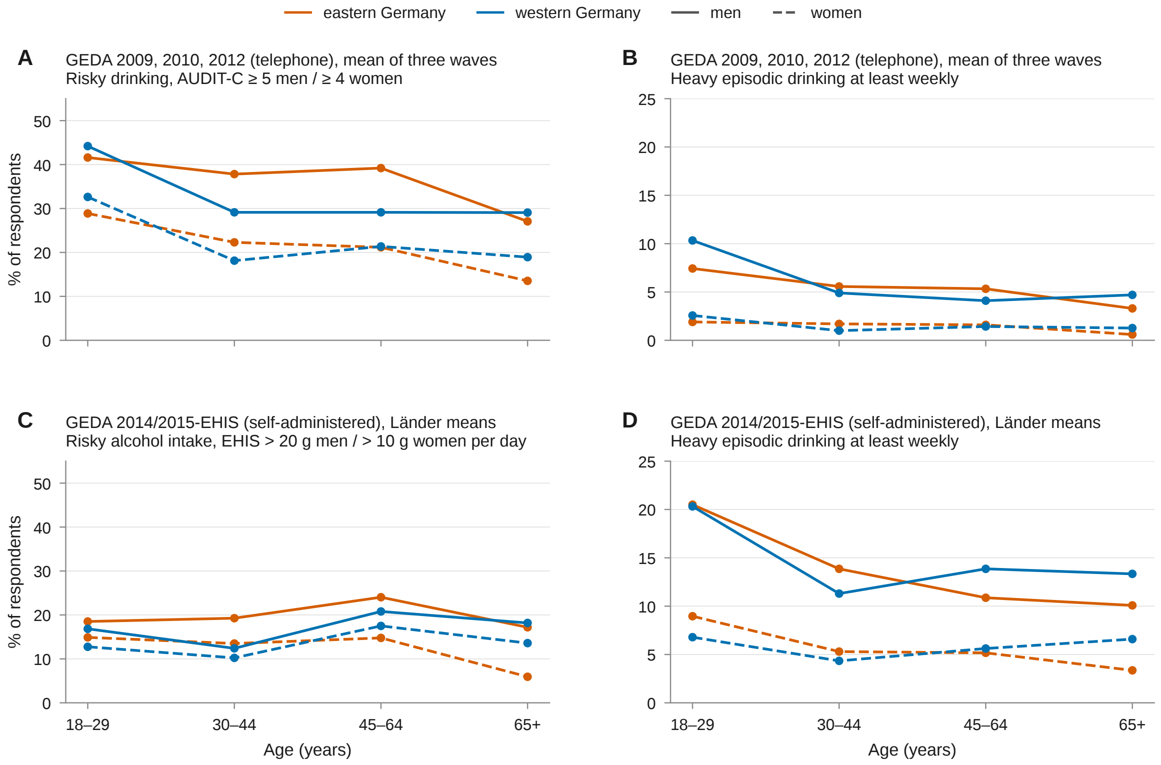


**Figure S4. Self-reported drinking pattern in eastern and western Germany by age, GEDA surveys.** Share of respondents by age band, eastern Germany (vermilion) and western Germany (blue), men (solid) and women (dashed). (A, B) Gesundheit in Deutschland aktuell 2009, 2010 and 2012 (telephone interviews, about 19 000 to 22 000 respondents per wave), shown as the mean of the three published estimates; eastern Germany here includes Berlin. (A) Risky drinking defined by an AUDIT-C score of at least 5 in men and 4 in women. (B) Six or more alcoholic drinks on one occasion at least weekly. (C, D) GEDA 2014/2015-EHIS (self-administered questionnaire, 24 016 respondents), published for the 16 Länder and aggregated as population-weighted means of the five eastern and the ten western Länder, Berlin excluded. (C) Risky alcohol intake defined by the EHIS frequency and quantity questions as more than 20 g (men) or 10 g (women) of pure alcohol per day among people who drink at least weekly. (D) Six or more drinks on one occasion at least weekly in the past 12 months. The two survey generations differ in instrument, answer categories, reference period and mode, so levels are compared between regions within a generation only. Source: GBE-Bund tables 192, 193, 26612 and 26613 (Robert Koch-Institut).

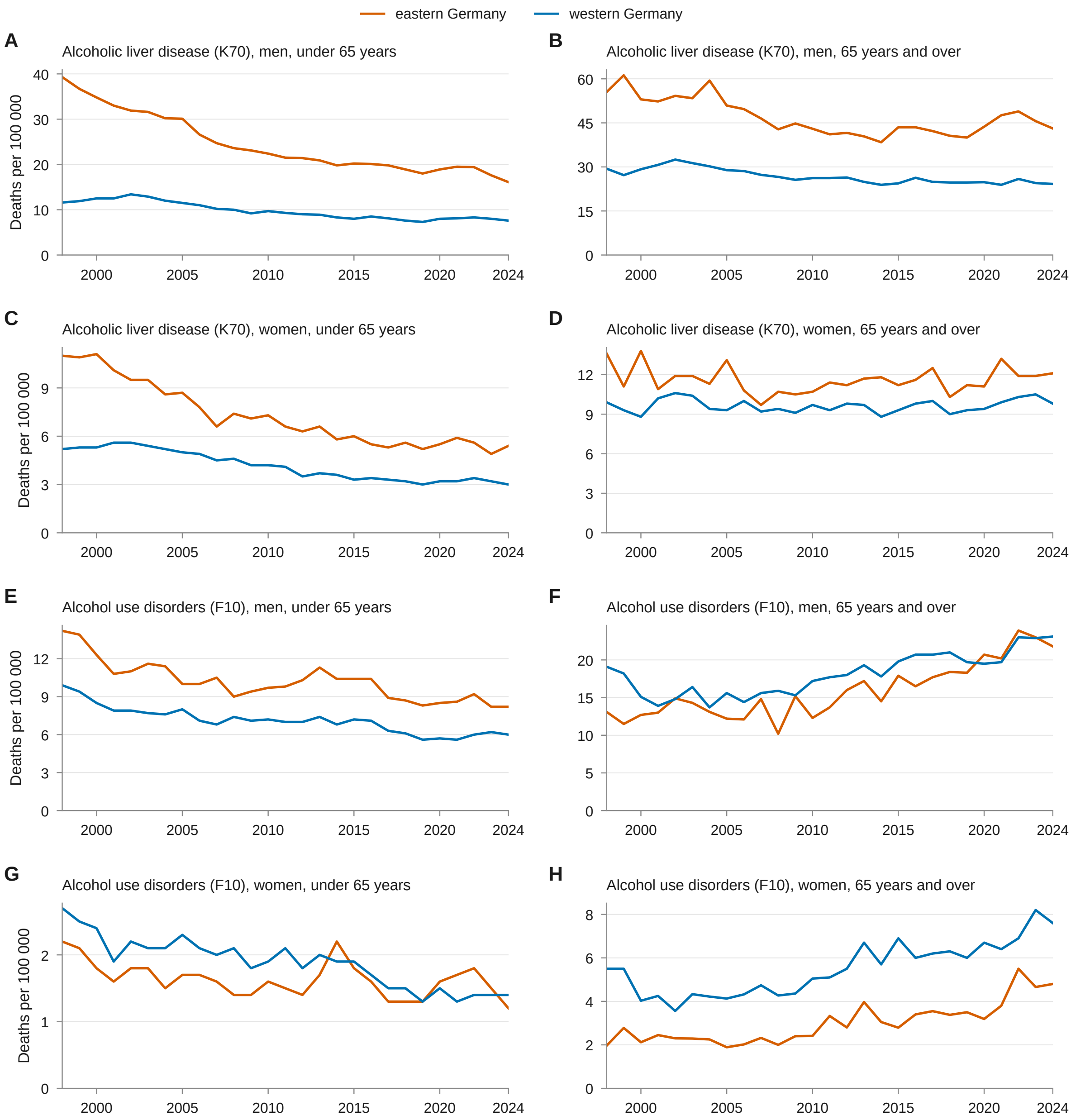


**Figure S5. Alcohol-specific mortality in eastern and western Germany by cause, sex and age band, 1998–2024.** Deaths from alcoholic liver disease (ICD-10 K70; A–D) and from mental and behavioural disorders due to alcohol (F10; E–H), eastern Germany (vermilion; new Länder without East Berlin) and western Germany (blue; former federal territory with East Berlin), men (A, B, E, F) and women (C, D, G, H), under 65 years (left column) and 65 years and over (right column), rates per 100 000 standardised to the European standard population 2013 within the age band. The y-axis scale differs between panels. The GBE-Bund table publishes the 100 most frequent causes of death per region, sex and age band; in years in which F10 was not among them for women aged 65 and over, the number of deaths was recovered as the difference between all ages and under 65 and the rate estimated by distributing these deaths over the 5-year age groups in proportion to the F10–F19 block of the full table (marked in the data file). Source: Statistisches Bundesamt, Todesursachenstatistik, via GBE-Bund (tables "Sterbefälle, Sterbeziffern, häufigste Todesursachen" and "Sterbefälle, Sterbeziffern (ab 1998)").
