## Supplementary material for "The plateau in liver disease mortality in Germany: contrasting trends by age, region and ICD-10 code, 2000–2024": STROBE checklist

### STROBE reporting checklist

Descriptive population-level mortality trend analysis | Manuscript and supplementary appendix

*Locations: M = manuscript page; A = appendix page. N/A = not applicable. Design-specific STROBE items are interpreted for repeated aggregate observations, not a followed cohort.*

| Topic | No. | STROBE recommendation | Where reported |
|---|---|---|---|
| TITLE AND ABSTRACT | | | |
| **Title and abstract** | **1** | (a) Indicate the study's design with a commonly used term in the title or the abstract | **M 1, 3** "Descriptive study" in Methods (p 3). |
| **Title and abstract** | **1** | (b) Provide in the abstract an informative and balanced summary of what was done and what was found | **M 1** Structured Summary reports sources, age-specific trends, regions and GBD comparison. |
| INTRODUCTION | | | |
| **Background/rationale** | **2** | Explain the scientific background and rationale for the investigation being reported | **M 2–3** Introduction describes mortality trends and alternative explanations. |
| **Objectives** | **3** | State specific objectives, including any prespecified hypotheses | **M 2–3** Study objectives stated; candidate explanations are not prespecified testable hypotheses. |
| METHODS | | | |
| **Study design** | **4** | Present key elements of study design early in the paper | **M 3** Methods opens with "This descriptive study" and identifies data sources. |
| **Setting** | **5** | Describe the setting, locations, and relevant dates, including periods of recruitment, exposure, follow-up, and data collection | **M 3–4; A 2–4, 8–9** Countries, regions, calendar years and source versions; no individual recruitment or follow-up. |
| **Participants** | **6** | (a) Cohort study—Give the eligibility criteria, and the sources and methods of selection of participants. Describe methods of follow-up<br>Case-control study—Give the eligibility criteria, and the sources and methods of case ascertainment and control selection. Give the rationale for the choice of cases and controls<br>Cross-sectional study—Give the eligibility criteria, and the sources and methods of selection of participants | **M 3–4; A 2–5** Units are registered deaths, hospital stays and resident populations; ICD and country selection described. |
| **Participants** | **6** | (b) Cohort study—For matched studies, give matching criteria and number of exposed and unexposed<br>Case-control study—For matched studies, give matching criteria and the number of controls per case | **N/A** No matching or individually recruited participants. |
| **Variables** | **7** | Clearly define all outcomes, exposures, predictors, potential confounders, and effect modifiers. Give diagnostic criteria, if applicable | **M 3–4; A 5–6, 8–9** ICD-10 outcomes, age/sex/region strata and exposure indicators defined; no confounder model. |
| **Data sources/ measurement** | **8*** | For each variable of interest, give sources of data and details of methods of assessment (measurement). Describe comparability of assessment methods if there is more than one group | **M 3–4; A 2–9** Sources, ICD definitions, survey instruments and comparability checks. |
| **Bias** | **9** | Describe any efforts to address potential sources of bias | **M 3–4, 10–11; A 5–9** Suppression/imputation sensitivity, small-count thresholds, coding and admission limitations. |

| Topic | No. | STROBE recommendation | Where reported |
|---|---|---|---|
| **Study size** | **10** | Explain how the study size was arrived at | **M 3–4** Administrative population-level series, not a recruited sample; nine age-band comparators selected for sufficient counts. |
| **Quantitative variables** | **11** | Explain how quantitative variables were handled in the analyses. If applicable, describe which groupings were chosen and why | **M 3–4; A 5–6, 8–9** Age bands, standard populations, three-year means and survey thresholds. |
| **Statistical methods** | **12** | (a) Describe all statistical methods, including those used to control for confounding | **M 3–4; A 5–8** Descriptive rates, standardisation and percent changes; no adjustment beyond age structure. |
| **Statistical methods** | **12** | (b) Describe any methods used to examine subgroups and interactions | **M 3–10** Stratified by sex, age, ICD code and geography; no interaction tests. |
| **Statistical methods** | **12** | (c) Explain how missing data were addressed | **M 3; A 5–6, 8–9** Unknown ages and suppressed cells imputed; F10 older-women rates estimated where absent. |
| **Statistical methods** | **12** | (d) Cohort study—If applicable, explain how loss to follow-up was addressed<br>Case-control study—If applicable, explain how matching of cases and controls was addressed<br>Cross-sectional study—If applicable, describe analytical methods taking account of sampling strategy | **N/A; A 8–9** No follow-up or matching; survey aggregates are published RKI estimates, not newly analysed microdata. |
| **Statistical methods** | **12** | (e) Describe any sensitivity analyses | **A 5–10; M 11** Alternative suppression allocation, GBD re-standardisation and F10 allocation validation. |
| **RESULTS** | | | |
| **Participants** | **13*** | (a) Report numbers of individuals at each stage of study—eg numbers potentially eligible, examined for eligibility, confirmed eligible, included in the study, completing follow-up, and analysed | **M 3–10; A 10–13** Different source coverages: Eurostat 36 countries, WHO 16, detailed age-band comparison 9; death/stay counts reported. |
| **Participants** | **13*** | (b) Give reasons for non-participation at each stage | **M 3, 5–6, 8–9; A 5–6** Restricted data availability and low counts determine which comparisons are interpretable; no participation process. |
| **Participants** | **13*** | (c) Consider use of a flow diagram | **N/A** No individual-level recruitment or inclusion flow; sources and subsets are explained in Methods. |
| **Descriptive data** | **14*** | (a) Give characteristics of study participants (eg demographic, clinical, social) and information on exposures and potential confounders | **M 4–10; A 10–13, 15–16** Rates and counts by age, sex, cause and region; population-level exposure summaries. |
| **Descriptive data** | **14*** | (b) Indicate number of participants with missing data for each variable of interest | **A 5–6, 8–11** Suppressed death cells and absent F10 rates documented; not individual-level missingness. |
| **Descriptive data** | **14*** | (c) Cohort study—Summarise follow-up time (eg, average and total amount) | **N/A** No individual follow-up. |
| **Outcome data** | **15*** | Cohort study—Report numbers of outcome events or summary measures over time | **N/A** No followed cohort. Population-level outcome trends are reported under the cross-sectional analogue below. |
| **Outcome data** | **15*** | Case-control study—Report numbers in each exposure category, or summary measures of exposure | **N/A** No case–control design. |

| Topic | No. | STROBE recommendation | Where reported |
|---|---|---|---|
| **Outcome data** | **15*** | Cross-sectional study—Report numbers of outcome events or summary measures | **M 4–10; A 10–12** Registered deaths, rates and hospital outcomes reported over calendar time; Table 1 and Figs 1–4. |
| **Main results** | **16** | (a) Give unadjusted estimates and, if applicable, confounder-adjusted estimates and their precision (eg, 95% confidence interval). Make clear which confounders were adjusted for and why they were included | **M 3–10; A 10–12** Age-standardised rates, counts and changes; published GBD 95% UIs. No multivariable estimates. |
| **Main results** | **16** | (b) Report category boundaries when continuous variables were categorized | **M 3; A 5–6, 8–9, 13** Age-band limits and drinking-risk cut-offs specified. |
| **Main results** | **16** | (c) If relevant, consider translating estimates of relative risk into absolute risk for a meaningful time period | **N/A** Absolute mortality and hospitalisation rates reported; no relative-risk estimates. |
| **Other analyses** | **17** | Report other analyses done—eg analyses of subgroups and interactions, and sensitivity analyses | **M 4–10; A 5–17** Sex/age/regional and GBD comparisons; sensitivity analyses documented in appendix. |
| **DISCUSSION** | | | |
| **Key results** | **18** | Summarise key results with reference to study objectives | **M 10** Opening Discussion summarises age, region, code and GBD findings. |
| **Limitations** | **19** | Discuss limitations of the study, taking into account sources of potential bias or imprecision. Discuss both direction and magnitude of any potential bias | **M 10–11; A 5–6, 8–9** Certification, imputation, outcome definitions, hospital admissions and missing lifetime exposure; quantified sensitivity where possible. |
| **Interpretation** | **20** | Give a cautious overall interpretation of results considering objectives, limitations, multiplicity of analyses, results from similar studies, and other relevant evidence | **M 10–11** Birth-cohort explanation qualified, alternatives and external evidence discussed; no causal attribution. |
| **Generalisability** | **21** | Discuss the generalisability (external validity) of the study results | **M 10–11; A 5–6, 8–9** National registry coverage; varying code definitions and certification constrain transfer to other settings. No formal external-validity analysis. |
| **OTHER INFORMATION** | | | |
| **Funding** | **22** | Give the source of funding and the role of the funders for the present study and, if applicable, for the original study on which the present article is based | **M 1, 4, 12** Summary: no funding. Role of funding source and declarations reported. |

* *For case–control studies, information should be reported separately for cases and controls; for cohort and cross-sectional studies, separately for relevant exposure groups where applicable.*
